# Reduced reciprocal inhibition during clinical tests of spasticity is associated with impaired reactive standing balance control in children with cerebral palsy

**DOI:** 10.1101/2023.11.07.23298160

**Authors:** Jente Willaert, Lena H. Ting, Anja Van Campenhout, Kaat Desloovere, Friedl De Groote

**Author notes:** **Corresponding author:** Jente Willaert, Gebouw De Nayer (bus 1501, lokaal 02.49), Tervuursevest 101, 3001 Leuven, Belgium.

## Abstract

**Background:** Joint hyper-resistance is a common symptom in cerebral palsy (CP). It is assessed by rotating the joint of a relaxed patient. Joint rotations also occur when perturbing functional movements. Therefore, joint hyper-resistance might contribute to reactive balance impairments in CP.

**Aim:** To investigate relationships between altered muscle responses to isolated joint rotations and perturbations of standing balance in children with CP.

*Methods & procedures:* 20 children with CP participated in the study. During an instrumented spasticity assessment, the ankle was rotated as fast as possible from maximal plantarflexion towards maximal dorsiflexion. Standing balance was perturbed by backward support-surface translations and toe-up support-surface rotations. Gastrocnemius, soleus, and tibialis anterior electromyography was measured. We quantified reduced reciprocal inhibition by plantarflexor-dorsiflexor co-activation and the neural response to stretch by average muscle activity. We evaluated the relation between muscle responses to ankle rotation and balance perturbations using linear mixed models.

*Outcomes & results:* Co-activation during isolated joint rotations and perturbations of standing balance was correlated across all levels. The neural response to stretch during isolated joint rotations and balance perturbations was not correlated.

*Conclusions & implications:* Reduced reciprocal inhibition during isolated joint rotations might be a predictor of altered reactive balance control strategies.

*Highlights:*

1. Impaired reciprocal inhibition might underlie altered balance control in CP.
2. Co-activation during isolated joint rotations and balance responses is correlated.
3. Hyperreflexia is not correlated with increased response to perturbations of standing.
4. Reduced reciprocal inhibition has functional implications.
5. It might be valuable to clinically assess reduced reciprocal inhibition.

*What this paper adds:* It has been hard to relate alterations in muscle coordination during functional movements to alterations in the muscle’s response to isolated joint rotations as applied during (clinical) assessments of hyper-reflexia. Here, we performed a more comprehensive assessment of the altered muscle response to isolated joint rotations in children with cerebral palsy (CP) by not only considering muscle activity in response to stretch but also agonist-antagonist co-activation. Muscle co-activation in response to isolated joint rotations in relaxed patients has been attributed to reduced reciprocal inhibition in the spinal cord. We found that muscle co-activation during isolated joint rotations was correlated to muscle co-activation during perturbed standing, an important functional movement. Therefore, increased muscle co-activation during standing balance control might – at least partially – result from reduced reciprocal inhibition in the spinal cord. In contrast, we found very few relations between the mean muscle activity during isolated joint rotations and perturbed standing. This might be due to the sensitivity of the response to stretch to stretch velocity, posture, and baseline muscle activity, all of which largely differed between the two conditions. Our results indicate that clinical assessment of reduced reciprocal inhibition during isolated joint rotations might provide information about balance impairments.

## INTRODUCTION

Joint hyper-resistance is the most common symptom in children with cerebral palsy (CP) (van den Noort et al., 2017). Joint hyper-resistance is clinically evaluated by assessing the resistance against an imposed passive muscle stretch (van den Noort et al., 2017). Muscle stretches also occur when functional movements are perturbed (e.g., when standing on a departing bus). Therefore, joint hyper-resistance might contribute to balance impairments that are common in children with CP (Pavão et al., 2013). Yet, little is known about the relation between joint hyper-resistance and balance impairments in CP.

Children with CP have higher muscle activity and higher levels of agonist-antagonist co-activation in response to perturbations of standing balance (Burtner et al., 1998; Willaert, Desloovere, et al., 2023; Willaert, Martino, et al., 2023). Children with CP activate their muscles more than typically developing (TD) children in response to similar center of mass (CoM) disturbances (Willaert, Desloovere, et al., 2023; Willaert, Martino, et al., 2023) and have higher muscle co-activation in response to both support-surface translations and rotations (Burtner et al., 1998; Willaert, Desloovere, et al., 2023; Willaert, Martino, et al., 2023).

Increased muscle co-activation observed in children with CP during perturbations of standing balance might be due to reduced reciprocal inhibition rather than being a compensation strategy to improve balance control. In an earlier study, we observed plantarflexor-dorsiflexor co-activation in response to both platform translations and rotations during standing in children with CP (Willaert, Desloovere, et al., 2023). Increased ankle stiffness due to increased muscle co-activation might help balance control in response to platform translations by resisting body movement with respect to the platform. However, increased joint stiffness does not help balance control during platform rotations, as it couples body motion with platform motion resulting in body tilt (Carpenter et al., 1999; Gollhofer, A et al., 1989; Horak & Nashner, 1986). A potential cause of the increased co-activation is reduced reciprocal inhibition, i.e., a lack of inhibition of the antagonistic muscle upon activation of the agonist (Crone C, 1993).

Differences in the response to perturbations of standing balance between children with CP and TD children, i.e., higher levels of muscle activity and co-activation, have striking similarities with differences in the response to isolated joint rotations in a relaxed condition. Muscle excitation in response to passive joint rotations is higher in children with CP than in TD children (Bar-On, Aertbeliën, et al., 2014; Dietz & Sinkjaer, 2007; Poon & Hui-Chan, 2009) and this increased muscle excitation is often attributed to spasticity or hyper-excitability of the stretch reflex (Dietz & Sinkjaer, 2007). Furthermore, isolated joint rotations elicit co-activation between the stretched muscle and its antagonist (e.g., gastrocnemius and tibialis anterior when rotating the ankle joint towards dorsiflexion) in children with CP, but not in TD children (Geertsen et al., 2018; Milner-Brown & Penn, 1979; Myklebust et al., 1982). This muscle co-activation has been attributed to reduced reciprocal inhibition (Leonard et al., 1990, 2006; Milner-Brown & Penn, 1979; Myklebust et al., 1982).

Neither current clinical scales nor instrumented tests of joint hyper-resistance assess reciprocal inhibition during isolated joint rotations in a relaxed patient (Milner-Brown & Penn, 1979; Myklebust et al., 1982). Clinical scales are commonly limited to the classification of the subjective feeling of the overall resistance to stretch by the examiner (Scholtes et al., 2006). During instrumented spasticity assessments, muscle activity is measured by electromyography (EMG), with the aim to distinguish neural and non-neural contributions to joint hyper-resistance (Bar-On et al., 2013; Sloot et al., 2017). Typically, the activity of the stretched muscle is reported as a measure of hyper-reflexia, while agonist-antagonist co-activation is not reported as a measure of altered reciprocal inhibition.

The failure to assess impairments in reciprocal inhibition might explain why no or only limited correlations have been found between the muscle response to isolated joint rotations and functional movement impairments (Bar-On, Molenaers, et al., 2014; Desloovere et al., 2006; Nielsen et al., 2020). Two prior studies investigated the relationship between joint hyper-resistance and reactive standing balance. These studies found no relationship between the Modified Ashworth Scale (MAS) and respectively CoM movement during standing in children with CP (Ali, 2021) and the ability to withstand perturbations without stepping in children with hereditary spastic paraparesis (de Niet et al., 2013). To our knowledge, the relationship between reduced reciprocal inhibition in response to isolated joint rotations and increased muscle co-activation during functional movements has not been studied.

We hypothesized that reduced reciprocal inhibition underlies plantarflexor-dorsiflexor co-activation in response to both isolated passive joint rotations and perturbations of standing in children with CP. Therefore, we expect that muscle co-activation during isolated joint rotations will be correlated to muscle co-activation during perturbations of standing balance. Secondary, we hypothesized that the neural response of the plantarflexors in response to isolated ankle dorsiflexion does not explain increased plantarflexor activity during perturbations of standing balance. Therefore, we do not expect that average muscle activity in response to isolated joint rotations correlates with average muscle activity during perturbations of standing balance.

## METHODS

### Participants

As there was no prior data, sample size was determined to enable detection of medium correlations (Spearman correlation of 0.4) with a power of 95%. The ethical committee of UZ/KU Leuven (S63321) approved this observational study. Twenty-one children with spastic CP participated in the study (table 1) between January 2021 and August 2021. Children were diagnosed by a neuro-pediatrician and met the following inclusion criteria: (1) aged 5 to 17 years; (2) Gross Motor Classification Scale (GMFCS) I-III; (3) able to stand independently for at least 10 minutes; (4) no orthopedic/neurological surgery or botulinum neurotoxin injections in respectively the previous 12 or 6 months. One child was excluded due to a lack of cooperation.

**Table 1:**
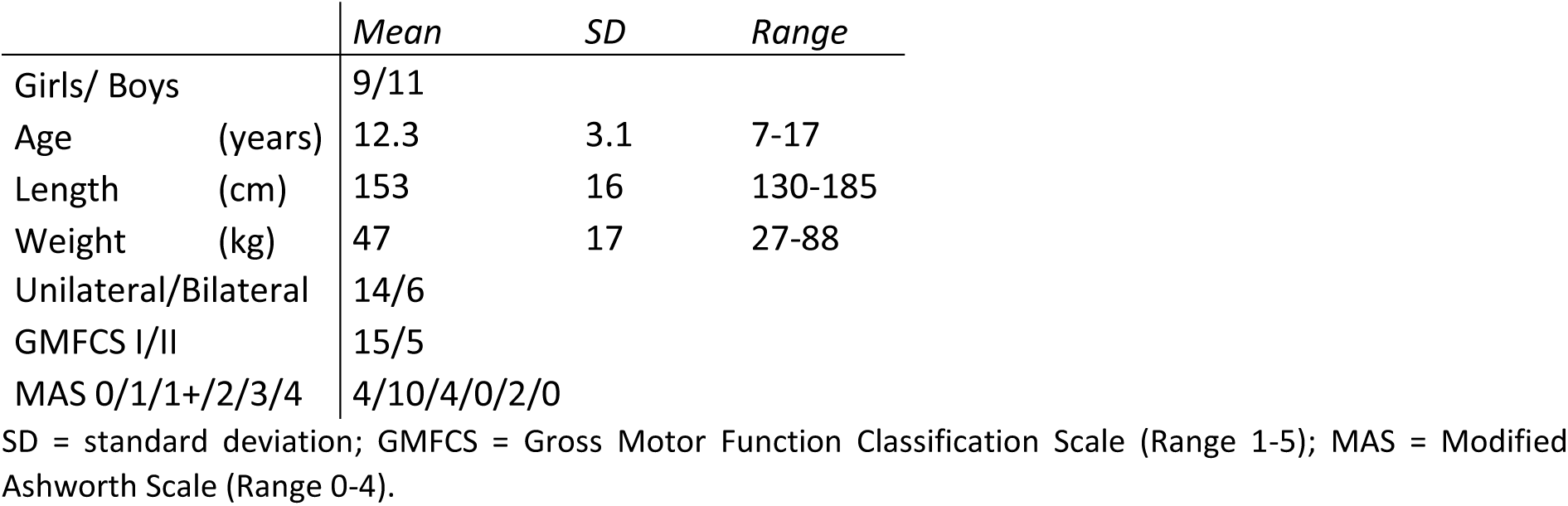
Demographic data of participants.

|  | *Mean* | *SD* | *Range* |
| --- | --- | --- | --- |
| Girls/ Boys | 9/11 |  |  |
| Age (years) | 12.3 | 3.1 | 7-17 |
| Length (cm) | 153 | 16 | 130-185 |
| Weight (kg) | 47 | 17 | 27-88 |
| Unilateral/Bilateral | 14/6 |  |  |
| GMFCS I/II | 15/5 |  |  |
| MAS 0/1/1+/2/3/4 | 4/10/4/0/2/0 |  |  |

SD = standard deviation; GMFCS = Gross Motor Function Classification Scale (Range 1-5); MAS = Modified Ashworth Scale (Range 0-4).

### Protocol

Children and their legal representative signed respectively informed assent and informed consent before the start of the measurements following the Declaration of Helsinki. All data was collected during a single session. The protocol consisted of a clinical assessment of range of motion and MAS; instrumented spasticity assessment of the plantarflexors, hamstrings and rectus femoris; and reactive balance assessments. Note that not all data was used in this study.

We performed an instrumented spasticity assessment of the plantarflexors by applying isolated joint rotations following a previously developed and reliable method (Bar-On et al., 2013; Schless et al., 2015). Participants lay supine and were asked to relax. The lower leg was supported by a customized frame that allowed ankle rotation. A researcher rotated the ankle joint as fast as possible (± 1s) from a plantar flexed position to the end range of motion towards dorsiflexion (figure 1, column 1). At least 7 seconds of rest were provided between each of the five trials to control for movement history dependence in muscle resistance to stretch (Lakie & Campbell, 2019; Willaert et al., 2020).

**Figure 1:**
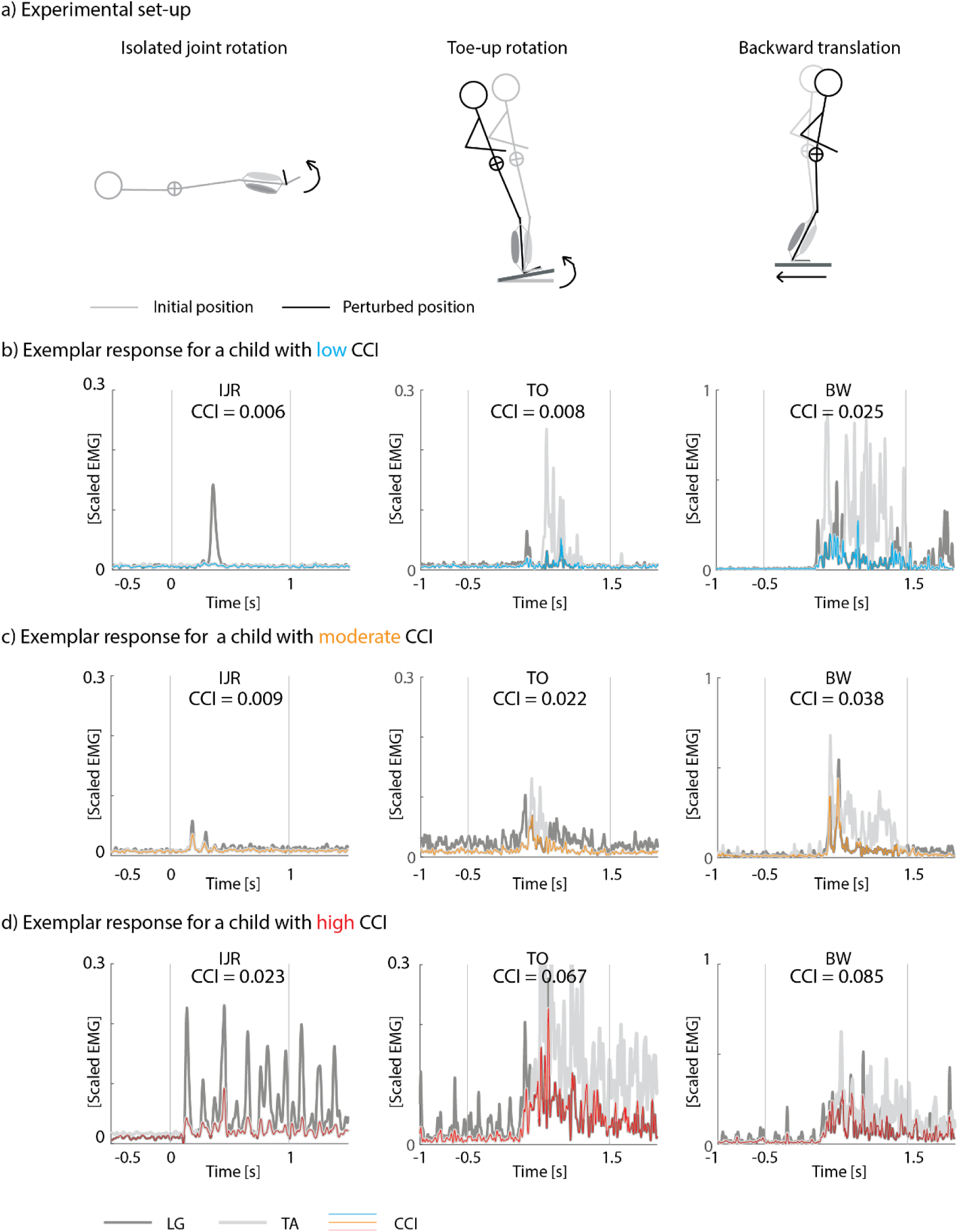
Experimental set-up (**a**) and exemplar responses (**b-d**) for the co-contraction index (CCI) during isolated joint rotations (IJR, column 1), toe-up rotations level 2 (TO, column 2), and backward translations level 2 (BW, column 3) for a child with low muscle co-activation (blue), moderate co-activation (orange), and high co-activation (red). Starting position of the experimental set-up in grey, perturbed position in black. The co-contraction index was calculated as the average of the minimum EMG signal of lateral gastrocnemius (LG, dark grey) and tibialis anterior (TA, light grey).

Muscle responses to perturbations of standing balance were measured on a Caren platform (Motek, Netherlands). Participants stood barefoot on the platform (starting position was marked and consistent between trials) and were secured using a safety harness. Instructions were to stand upright and to maintain balance without stepping unless necessary to avoid falling. The perturbation protocol consisted of (1) backward translations (figure 1, column 3), followed by (2) toe-up rotations (figure 1, column 2). The plantarflexors are stretched by both backward translations and toe-up rotations but since backward translations cause a forward rotation of the body and toe-up rotations cause a backward rotation of the body, plantarflexor activity elicited by muscle stretch will only aid in maintaining an upright posture in response to translational perturbations. In addition, increased ankle stiffness due to muscle co-activation will only aid to stay upright in response to translational perturbations but not in response to rotational perturbations. We applied ten series of eight identical perturbation trials starting with six increasingly difficult translational perturbation levels (increasing platform displacement, velocity, and/or acceleration) followed by four increasingly difficult rotational perturbation levels (details on platform movement are described in Supplementary material S1, figure S1). When the participant stepped in more than three trials within one level, we did not continue to the next level (Willaert, Desloovere, et al., 2023; Willaert, Martino, et al., 2023). If needed, rest was given between levels.

Muscle activity of the lateral gastrocnemius (LG), medial gastrocnemius (MG), soleus (SOL), and tibialis anterior (TA) was measured using surface electromyography (EMG) at 1000Hz (ZeroWire EMG Aurion, Cometa, Italy). Electrodes (Ambu Blue Sensor, Ballerup, Denmark) were placed according to SENIAM guidelines (Hermens et al., 2000).

### Data processing & analysis

EMG data was filtered using a fourth order Butterworth band-pass filter with 10 and 450Hz cut-offs, rectified, and low-pass filtered with a fourth order Butterworth filter with 40Hz cut-off. The filtered EMG signal was scaled to the maximum value observed across all movements performed during the protocol (i.e., maximum voluntary contractions for plantarflexors and TA, isolated joint rotations for ankle and knee, perturbations of standing, squats, and jumps). For the isolated joint rotations, average scaled muscle activity was calculated across all five trials for each participant. For the perturbations of standing, average scaled muscle activity was calculated across all non-stepping trials within one level for each participant.

### Outcome parameters

We computed the **co-contraction index (CCI)**, a measure of muscle co-activation, as the minimum of TA and respectively LG, MG, and SOL filtered and scaled EMG averaged over the time interval of interest, which is proportional to the common area under the EMG trajectories (Frost et al., 1997) (equation 1) (figure 1).

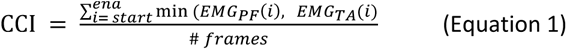

With PF referring to LG, MG, or SOL; *i* referring to the different time frames, and # frames the length of the analyzed time period with *start* the first frame and *stop* the last frame. For the isolated joint rotations, we analyzed the EMG signal over 1s following rotation onset. For the perturbations of standing balance, we analyzed the EMG signal from 0.5s before until 1.5s after onset.

We assessed **mean muscle activity** in response to stretch as the time-averaged processed and scaled EMG for all muscles individually using the same time periods as for the CCI. During isolated joint rotations, the mean muscle activity is a measure of the neural response to stretch consisting of both short-latency stretch reflexes and prolonged muscle activity in response to stretch that is often observed in CP (Bar-On, Aertbeliën, et al., 2014). During reactive balance, the mean muscle activity captures the neural response to stretch as well as balance correcting responses.

The most affected leg (based on MAS) was analyzed.

### Statistical analysis

All statistical analysis were performed using Matlab (2018, Mathworks, United States) with differences considered significant at p<0.05.

We explored correlations between isolated joint rotations and the different reactive balance tasks (different conditions and levels) for (1) muscle co-activation (CCI) and (2) mean muscle activity using Spearman correlations, as our data did not follow a normal distribution. No corrections for multiple testing were performed as the aim was to explore possible relationships.

In addition, we assessed the relation between the muscle response to isolated joint rotations and to translational or rotational perturbations across perturbations levels using mixed linear models (equation 2). Independent variables were perturbation level and the muscle response (CCI or mean activity) during isolated joint rotations and the dependent variable was the muscle response during the reactive balance conditions. A participant factor (i.e., subject in equation 2) was included as random factor nested within group.

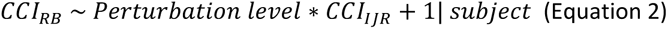

With *CCI*_*RB*_ the co-contraction index for toe-up rotational perturbations or backward translational perturbations, and *CCI*_*IJR*_the co-contraction index for the isolated joint rotations.

We created different models for the different outcomes (CCI and mean muscle activity), different muscle pairs/muscles, and different standing balance perturbations (translations or rotations).

## RESULTS

Due to empty EMG batteries, we had to exclude SOL data for one child and TA data for another child.

### Standing balance performance

For toe-up rotations, all children performed level 1 but respectively four and one children did not perform levels 2-4 and levels 3-4. For backward translations, all children performed level 1 but respectively two, one, two, and five children did not perform levels 2-6, levels 3-6, levels 4-6, and levels 5-6 (supplementary material S2, table S1).

### Muscle co-activation & mean muscle activity

We observed a large inter-subject variability in co-contraction index values and mean muscle activity values for the isolated joint rotations, toe-up rotations, and backward translations. Yet, our sample covered the complete range without clear outliers (figure 2, Supplementary material S3, table S2-3, figure S2-S3).

**Figure 2:**
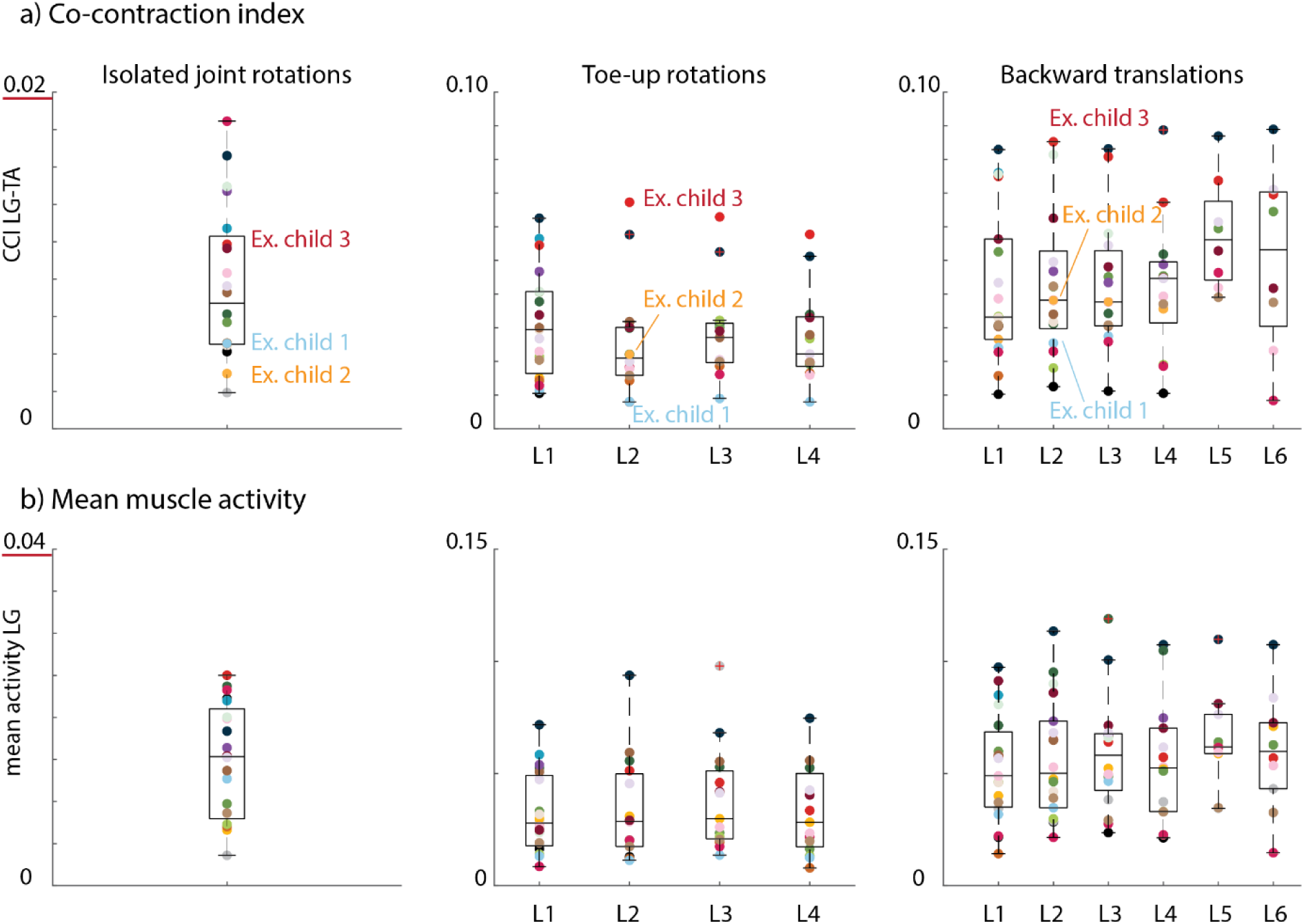
Co-contraction index for LG-TA **(a)** and mean muscle activity for LG **(b)** across all participants for isolated joint rotations (left column), toe-up rotations (middle column), and backward translations (right column). Every dot represents one child. L1-L6 = Levels 1 to 6.

### Relation between muscle co-activation during isolated joint rotations and perturbed standing

The CCI during isolated joint rotations was correlated to the CCI during perturbations of standing balance for some muscle pairs and conditions. Overall, we found most correlations for LG-TA and rotational perturbations (figure 3-4 and supplementary material S4, table S4). LG-TA co-activation during isolated joint rotations was correlated with LG-TA co-activation during perturbed standing for the lowest two levels of toe-up rotations and backward translations (figure 3-4). MG-TA co-activation during isolated joint rotations was correlated with MG-TA co-activation during perturbed standing for toe-up rotations of level 2 and 4 (figure 4, figure S4). SOL-TA co-activation during isolated joint rotations was related with SOL-TA co-activation during perturbed standing for toe-up rotations of level 2 (figure 4, figure S5).

**Figure 3:**
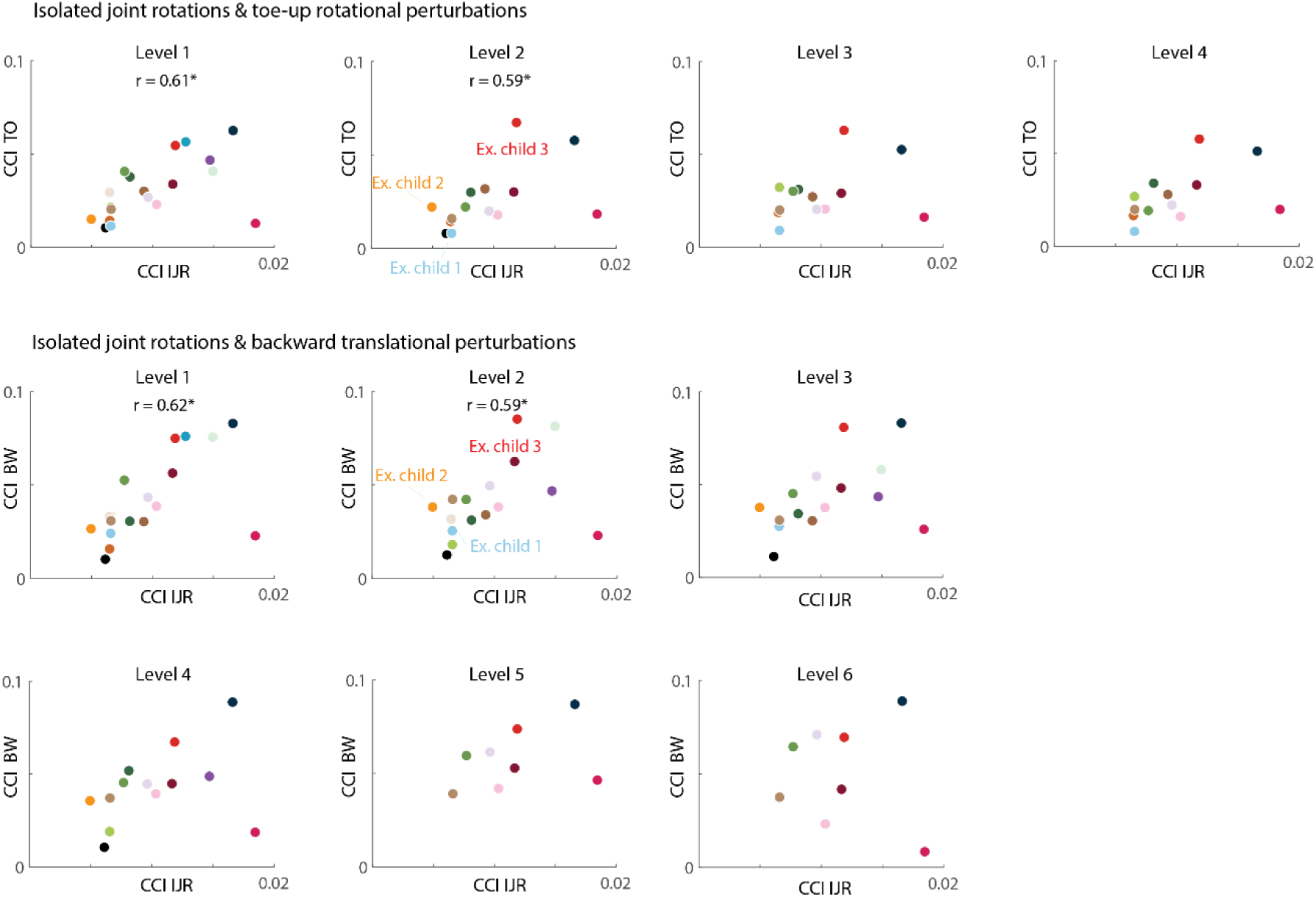
Spearman correlations between lateral gastrocnemius (LG) and tibialis anterior (TA) co-activation (CCI) during isolated joint rotations (IJR) and during perturbations of standing balance (toe-up rotations (TO): upper part; backward translations (BW): lower part) for each level. Each dot represents one child. Significant associations (p<0.05) are indicated with a star and corresponding r value.

**Figure 4:**
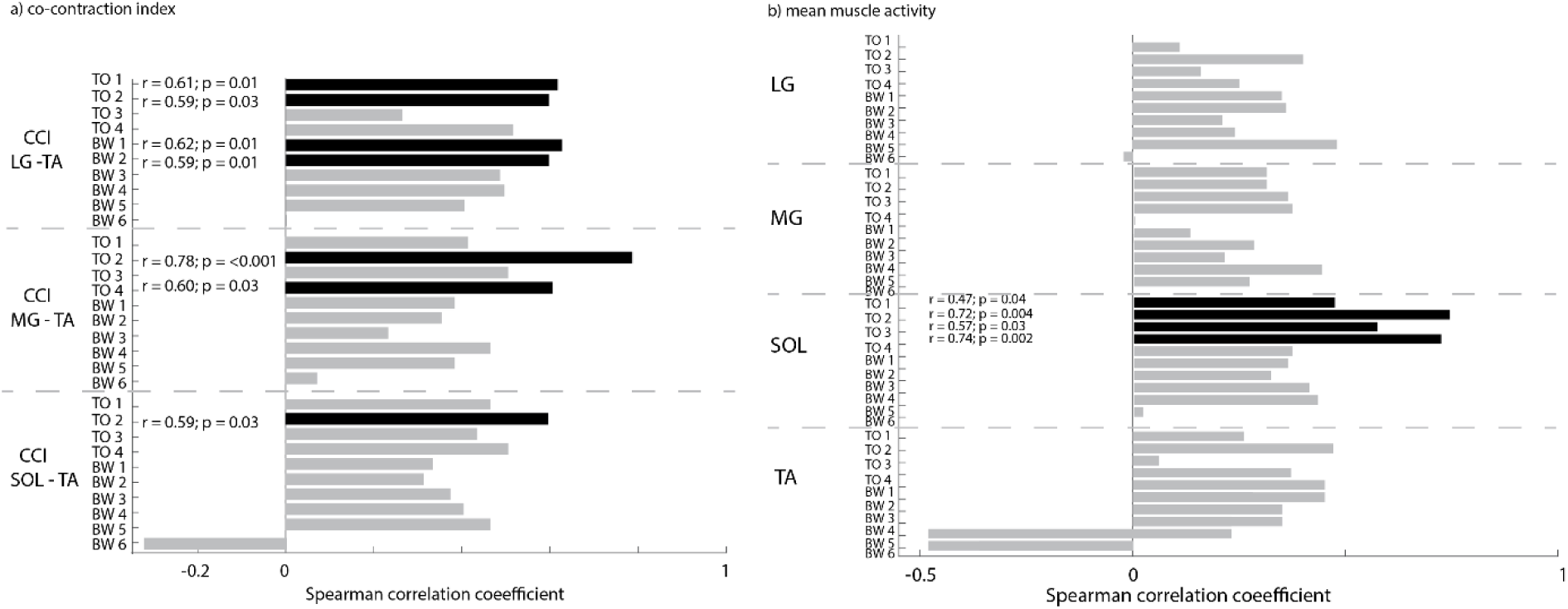
Associations (Spearman r coefficient and p-values) between the co-contraction index (CCI) (left) and mean muscle activity (right) during isolated joint rotations and perturbations of standing balance (TO = toe-up rotations; BW = backward translations). Significant relations are indicated in black. LG = lateral gastrocnemius; MG =medial gastrocnemius; SOL = soleus; TA = tibialis anterior.

Our analysis based on linear mixed models revealed a relation between co-activation during perturbations of standing balance and during isolated joint rotations for toe-up rotations for all muscle pairs and backward translations for LG-TA and MG-TA.

We found a significant main effect of LG-TA CCI during isolated joint rotations on LG-TA CCI during toe-up rotations (p=0.006) and backward translations (p=0.004). No main effect of level was found. No interaction effect between level and CCI during isolated joint rotations was found for toe-up rotations, while there was an interaction effect for backward translations (p=0.025) (supplementary material S5, table S5), suggesting a different relation for level 6.

We also found a significant main effect of MG-TA CCI during isolated joint rotations on MG-TA CCI during toe-up rotations (p=0.003) and backward translations (p=0.024). No main effect of level or interaction effect was found for toe-up rotations. For backward translations, there was a main effect for level (p=0.002), suggesting higher levels of co-activation with increasing perturbation level, and an interaction effect for level and co-activation during isolated joint rotations (p=0.001) (supplementary material S5, table S5).

We found a significant main effect of SOL-TA CCI during isolated joint rotations on SOL-TA CCI during toe-up rotations (p=0.047) but not for backward translations. No significant main effects for level or interaction effects were found (supplementary material S5, table S5).

### Relation between muscle activity during isolated joint rotations and perturbed standing

We only found associations between mean muscle activity during isolated joint rotations and mean muscle activity during toe-up perturbations of standing for the soleus (p<0.03) (figure 4 and supplementary material S6, table S6, figure S6-S9).

Similarly, based on the linear mixed model, we found that muscle activity during isolated joint rotations was only related to muscle activity during perturbed standing for the soleus for toe-up rotations and not backward translations. We found a significant main effect of mean SOL activity during isolated joint rotations on mean SOL activity during toe-up rotations (p=0.034). Further, we found an interaction effect for level and mean activity during isolated joint rotations for LG during toe-up rotations (p=0.033) and backward translations (p=0.005) and for the SOL during toe-up rotations (p=0.005) (supplementary material S7, table S7).

## DISCUSSION

Our results suggest that reduced reciprocal inhibition contributes to altered reactive balance control in children with CP. We found that muscle co-activation in response to isolated joint rotations was related to increased muscle co-activation in response to perturbations of standing balance in children with CP. Muscle co-activation in response to isolated joint rotations in relaxed patients has been attributed to reduced reciprocal inhibition in the spinal cord given the absence of other control processes in this condition (Leonard et al., 1990). The observed correlations thus suggest that the increased muscle co-activation during standing balance control might at least partially rely in spinal processes. In contrast, we found very few relations between the mean muscle activity during isolated joint rotations and perturbed standing. Current assessment of joint hyper-resistance focuses on reflex hyper-excitability and alterations in passive tissue properties (Bar-On et al., 2013) but our results indicate that clinical assessment of reduced reciprocal inhibition during isolated joint rotations might provide information about balance impairments.

In contrast to many previous studies, we found a relation between the response to muscle stretch at rest and functional movements. Previous research has mainly focused on the relation between spasticity or reflex hyper-excitability and muscle activity during walking (Nielsen et al., 2020). Only two prior studies specifically investigated reactive standing balance and found no relation between joint hyper-resistance as measured by MAS and CoM movement (Ali, 2021) or the ability to withstand perturbations without stepping (de Niet et al., 2013). In contrast to prior studies, we did not only assess general resistance or the neural response to muscle stretch but also muscle co-activation, as a sign of reduced reciprocal inhibition. We specifically related those outcomes to muscle coordination during perturbed standing. This allowed us to relate observations at rest to observations during a functional task. An exploratory analysis showed that the MAS score was not correlated to muscle co-activation during reactive standing (supplementary material S9, table S10-S11, figures S10-S12), confirming prior work and stressing the importance of using more specific outcomes (Scholtes et al., 2006). Alterations in passive tissue properties such as contractures might also contribute to functional impairments (Nielsen et al., 2020) but an exploratory analysis showed that co-activation during balance perturbations was not related to passive joint stiffness, suggesting that mechanical tissue properties do not explain variability in muscle coordination underlying balance control (supplementary material S10, table S12).

The associations between muscle co-activation during isolated joint rotations and during reactive standing suggest a common neural deficit. Both spinal and supraspinal pathways are involved in reactive standing balance whereas the response to isolated joint rotations is mainly driven by spinal pathways (Dietz & Sinkjaer, 2007). Hence, associations in CCI between conditions suggest that reactive balance impairments in children with CP might at least partially originate from deficits in spinal pathways. However, we cannot exclude contributions from reduced selective control, i.e., common drive to agonists and antagonists due to impaired corticospinal tracts. We found associations in muscle co-activation between isolated joint rotations and perturbed standing notwithstanding striking differences in baseline muscle activity, body position, and stretch velocity. In both conditions, the ankle was dorsiflexed by an external force. During isolated joint rotations, the patient is at rest and stretch velocity is high whereas during perturbed standing, muscles are active, and the stretch velocity is lower (factor ten). Hence, reduced reciprocal inhibition might be consistent across stretch velocities and tasks. In contrast, the stretch reflex is known to be velocity- and task-dependent (Bar-On, Molenaers, et al., 2014; Carpenter et al., 1999; Dietz & Sinkjaer, 2007), which might explain the limited associations between responsive muscle activity between conditions. It would be interesting to investigate whether associations between response muscle activity in response to isolated joint rotations and perturbations of standing balance are present for balance perturbations with higher accelerations, velocities, or displacements.

Although increased muscle co-activation might hinder balance control, we did not observe a relation between muscle co-activation and balance performance. Visual inspection of our data suggests that muscle co-activation is not related to the ability to withstand perturbations without stepping (figure 3, figure S2-S3), suggesting that other factors might be important here. Fear of falling might have induced stepping as stepping is an effective way to increase the base of support. Muscle co-activation was also not related to CoM movement during non-stepping responses (supplementary material S8, table S8-S9), suggesting that children compensate for the higher antagonistic muscle activity by also increasing agonistic muscle activity (Willaert, Desloovere, et al., 2023; Willaert, Martino, et al., 2023). Whereas such compensation strategies might be effective for small perturbation magnitudes, they are limited as activation bounds will be reached sooner when co-activating antagonistic muscles. In the future, it would be interesting to investigate whether reduced reciprocal inhibition is related to falling.

We did not explore relations between muscle responses to isolated joint rotations and perturbations of standing balance in TD children given that the muscle response to isolated joint rotations was small in these children. In addition, we previously showed that TD children have little muscle co-activation in response to translational and rotational perturbations of standing balance. We provide reference data for CCI and mean muscle activity levels across conditions for TD children to allow the reader to judge which part of the response might be deemed CP-specific (supplementary material S11, figure S13-15).

It is unlikely that our EMG scaling method affected our primary outcomes. EMG was scaled to the maximal signal across different tasks, and it is possible that not all children activated their muscles to the same extent. A higher scaling factor (due to higher activation during one of the tasks) would result in a smaller scaled signal and thus smaller mean muscle activity and CCI for both isolated joint rotations and perturbed standing balance. The absence of correlations between mean muscle activity during isolated joint rotations and during perturbed standing for most muscles, conditions, and levels thus suggests that the observations of the current study were not caused by inter-subject differences in scaling. In addition, we obtained similar results when scaling the EMG to the maximal signal across perturbation trials only or scaling the EMG to the maximal signal observed during translational and rotational perturbation of level 1 (performed by all subjects). Also, we chose a specific outcome measure for co-activation that is sensitive to both the presence and amplitude of the common activation but did not capture whether activation patterns, including timing of activation, were similar between conditions. Whereas evaluating the timing of muscle activation could provide additional information, such analysis would have been hard during reactive standing balance due to the presence of both balance corrective muscle activity and antagonistic activity in both plantarflexors and dorsiflexors.

## Conclusion

We demonstrated that muscle co-activation during isolated joint rotations and perturbations of standing balance is related. This suggests that reduced reciprocal inhibition, which is typically assessed during isolated joint rotations, might contribute to muscle co-activation during functional movements but this should be further investigated in other tasks.

## List of abbreviations

CP: Cerebral palsy
TD: Typically developing
MAS: Modified Ashworth Score
CoM: Center of mass
GMFCS: Gross motor function classification scale
EMG: Electromyography
LG: Lateral gastrocnemius
MG: Medial gastrocnemius
SOL: Soleus
TA: Tibialis anterior
CCI: Co-contraction index
IJR: Isolated joint rotation
TO: Toe-up rotation
BW: Backward translation

## Data availability

The data supporting the conclusions of this article will be made available by the authors upon request, without undue reservation.

## Author contributions

**Jente Willaert:** Funding acquisition, Data curation, Formal analysis, Investigation, Methodology, Project administration, Software, Validation, Visualization, Writing original draft

Lena H. Ting: Funding acquisition, Supervision, Visualization, Writing review & editing

Anja Van Campenhout: Resources, Supervision, Writing review & editing

Kaat Desloovere: Funding acquisition, Methodology, Resources, Supervision, Writing review & editing

Friedl De Groote: Funding acquisition, Conceptualization, Investigation, Methodology, Project administration, Resources, Supervision, Visualization, Writing review & editing

## Acknowledgements

There are no conflicts of interest.

This study was funded by the Flemish Agency for Scientific Research (FWO-Vlaanderen) through a research fellowship to JW (1192320N) and by the National Institute of Health (NIH) research grant NIH R01 HD46922 to LHT.

