## Supplementary Material for "Reduced reciprocal inhibition during clinical tests of spasticity is associated with impaired reactive standing balance control in children with cerebral palsy"

**S1. Perturbation profiles**

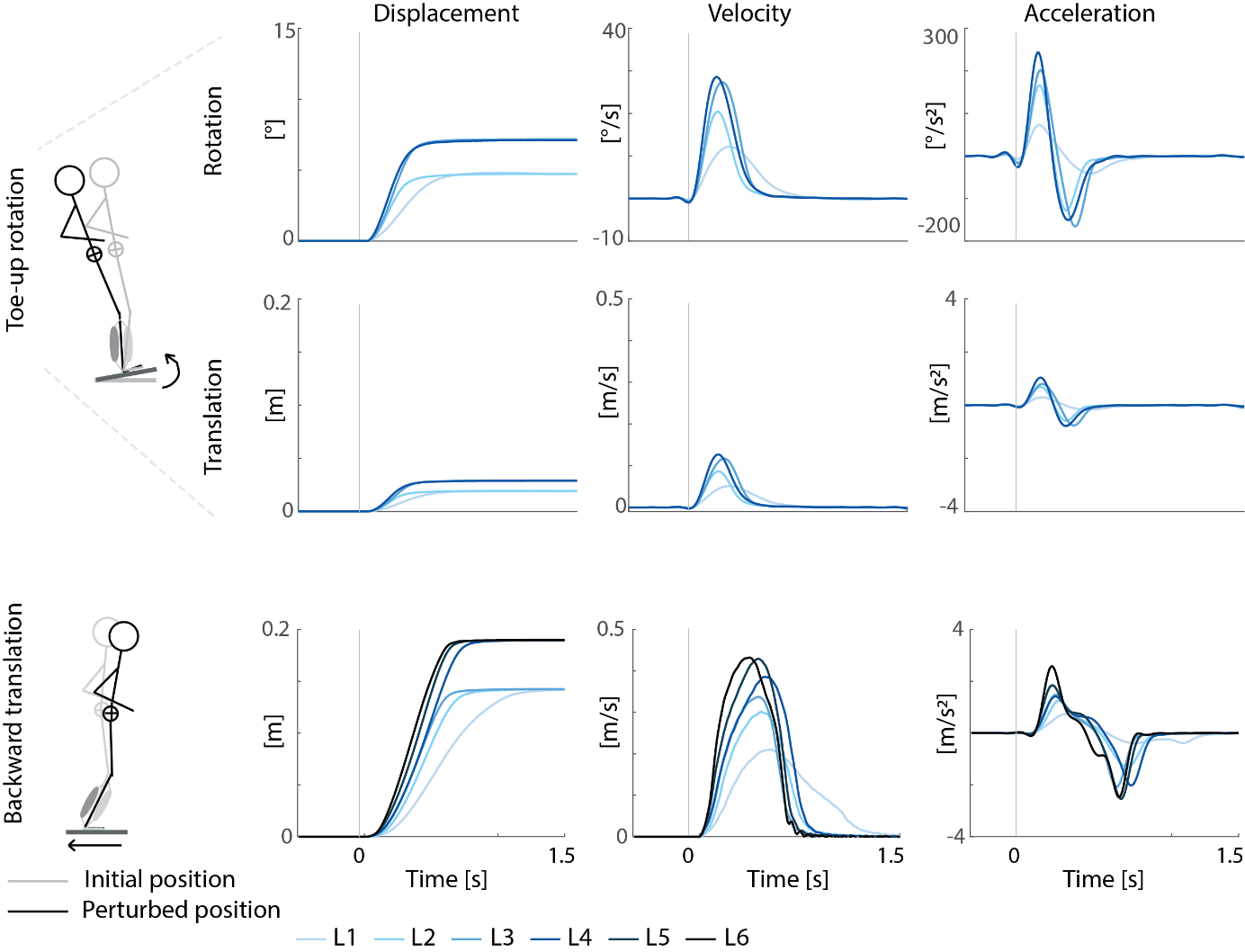

Figure S1: Toe-up rotational and backward translational perturbation profiles for the different levels.

**S2. Performed perturbation levels**

Table S1: Performed perturbation levels for all participants.

|  | **Toe-up rotations** | | | | **Backward translations** | | | | | |
| --- | --- | --- | --- | --- | --- | --- | --- | --- | --- | --- |
|  | L1 | L2 | L3 | L4 | L1 | L2 | L3 | L4 | L5 | L6 |
| CP1 | ✓ | ✓ |  |  | ✓ | ✓ | ✓ | ✓ |  |  |
| CP2 | ✓ | ✓ | ✓ | ✓ | ✓ | ✓ | ✓ | ✓ | ✓ | ✓ |
| CP3 | ✓ | ✓ | ✓ | ✓ | ✓ | ✓ | ✓ | ✓ | ✓ | ✓ |
| CP4 | ✓ | ✓ | ✓ | ✓ | ✓ |  |  |  |  |  |
| CP5 | ✓ |  |  |  | ✓ |  |  |  |  |  |
| CP6 | ✓ | ✓ | ✓ | ✓ | ✓ | ✓ | ✓ | ✓ | ✓ | ✓ |
| CP7 | ✓ | ✓ | ✓ | ✓ | ✓ | ✓ | ✓ | ✓ | ✓ | ✓ |
| CP8 | ✓ | ✓ | ✓ | ✓ | ✓ | ✓ | ✓ | ✓ |  |  |
| CP9 | ✓ | ✓ | ✓ | ✓ | ✓ | ✓ | ✓ | ✓ | ✓ | ✓ |
| CP10 | ✓ | ✓ | ✓ | ✓ | ✓ | ✓ | ✓ | ✓ |  |  |
| CP11 | ✓ |  |  |  | ✓ | ✓ |  |  |  |  |
| CP12 | ✓ | ✓ | ✓ | ✓ | ✓ | ✓ | ✓ |  |  |  |
| CP13 | ✓ | ✓ | ✓ | ✓ | ✓ | ✓ | ✓ | ✓ |  |  |
| CP14 | ✓ |  |  |  | ✓ | ✓ | ✓ | ✓ |  |  |
| CP15 | ✓ | ✓ | ✓ | ✓ | ✓ | ✓ | ✓ | ✓ | ✓ | ✓ |
| CP16 | ✓ | ✓ | ✓ | ✓ | ✓ | ✓ | ✓ | ✓ | ✓ | ✓ |
| CP17 | ✓ |  |  |  | ✓ | ✓ | ✓ |  |  |  |
| CP18 | ✓ | ✓ | ✓ | ✓ | ✓ | ✓ | ✓ | ✓ | ✓ | ✓ |
| CP19 | ✓ | ✓ | ✓ | ✓ | ✓ | ✓ | ✓ | ✓ | ✓ | ✓ |
| CP20 | ✓ | ✓ | ✓ | ✓ | ✓ | ✓ | ✓ | ✓ | ✓ | ✓ |

L1-L6 indicate perturbation levels.

**S3. Mean and range of CCI and mean muscle activity.**

Table S2: Co-contraction index for all muscle pairs during isolated joint rotations and perturbations of standing balance.

|  | **LG-TA** | | **MG-TA** | | **SOL-TA** | |
| --- | --- | --- | --- | --- | --- | --- |
|  | *Median* | *Range* | *Median* | *Range* | *Median* | *Range* |
| **IJR** | 0.009 | 0.004 – 0.019 | 0.008 | 0.002 – 0.023 | 0.010 | 0.003 – 0.022 |
| **TO** | 0.023 | 0.008 – 0.067 | 0.017 | 0.007 – 0.076 | 0.024 | 0.010 – 0.089 |
| **BW** | 0.042 | 0.008 – 0.116 | 0.042 | 0.007 – 0.126 | 0.045 | 0.009 – 0.095 |

IJR = Isolated joint rotation; TO = toe-up rotational perturbation; BW = backward translational perturbation; LG = lateral gastrocnemius; MG = medial gastrocnemius; SOL = soleus; TA = tibialis anterior.

Table S3: Mean muscle activity for all muscles during isolated joint rotations and perturbations of standing balance.

|  | **LG** | | **MG** | | **SOL** | | **TA** | |
| --- | --- | --- | --- | --- | --- | --- | --- | --- |
|  | *Median* | *Range* | *Median* | *Range* | *Median* | *Range* | *Median* | *Range* |
| IJR | 0.015 | 0.004 - 0.025 | 0.016 | 0.001 - 0.109 | 0.014 | 0.003 - 0.037 | 0.011 | 0.005 - 0.041 |
| TO | 0.029 | 0.008 - 0.165 | 0.021 | 0.005 - 0.154 | 0.031 | 0.009 - 0.073 | 0.036 | 0.010 - 0.128 |
| BW | 0.057 | 0.014 - 0.119 | 0.052 | 0.009 - 0.194 | 0.066 | 0.020 - 0.126 | 0.061 | 0.010 - 0.144 |

IJR = Isolated joint rotation; TO = toe-up rotational perturbation; BW = backward translational perturbation; LG = lateral gastrocnemius; MG = medial gastrocnemius; SOL = soleus; TA = tibialis anterior.

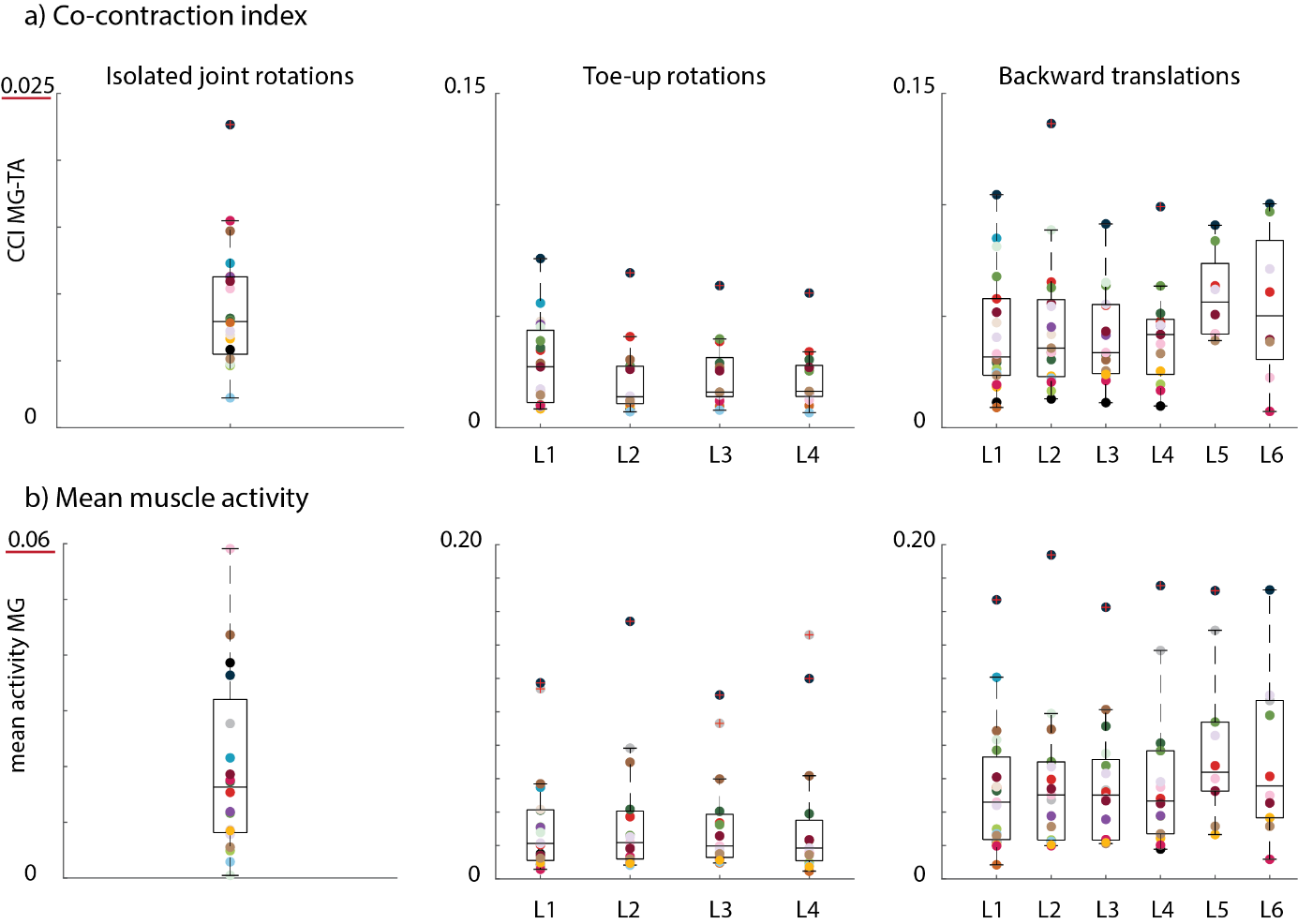

Figure S2: Co-contraction index for MG-TA **(a)** and mean muscle activity for MG **(b)** across all participants for isolated joint rotations (left column), toe-up rotations (middle column), and backward translations (right column). Every dot represents one child. L1-L6 = Levels 1 to 6.

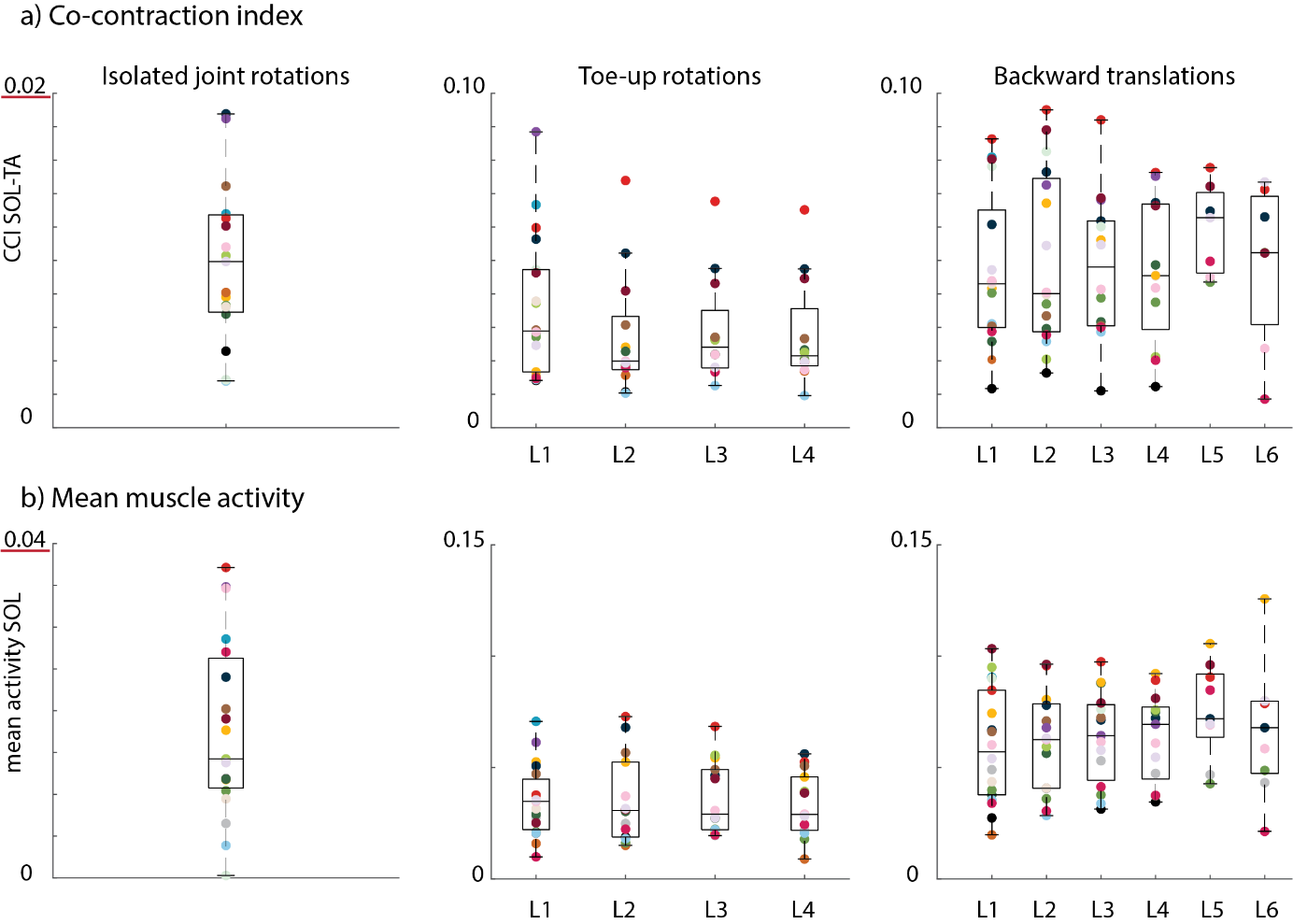

Figure S3: Co-contraction index for SOL-TA **(a)** and mean muscle activity for SOL **(b)** across all participants for isolated joint rotations (left column), toe-up rotations (middle column), and backward translations (right column). Every dot represents one child. L1-L6 = Levels 1 to 6.

**S4. Associations for the co-contraction index during isolated joint rotations and perturbations of standing balance.**

Table S4: Associations (r and p-values) for muscle co-activation during isolated joint rotations and perturbations of standing balance.

|  |  | **Toe-up rotations** | | | | **Backward translations** | | | | | |
| --- | --- | --- | --- | --- | --- | --- | --- | --- | --- | --- | --- |
|  |  | **L1** | **L2** | **L3** | **L4** | **L1** | **L2** | **L3** | **L4** | **L5** | **L6** |
| LG & TA | *r* | **0.61** | **0.59** | 0.26 | 0.51 | **0.62** | **0.59** | 0.48 | 0.49 | 0.40 | 0.00 |
|  | *p* | **0.01** | **0.03** | 0.38 | 0.07 | **0.01** | **0.01** | 0.07 | 0.09 | 0.33 | 1.00 |
| MG & TA | *r* | 0.41 | **0.78** | 0.50 | **0.60** | 0.38 | 0.35 | 0.23 | 0.46 | 0.38 | 0.07 |
|  | *p* | 0.08 | **< 0.001** | 0.08 | **0.03** | 0.11 | 0.17 | 0.42 | 0.12 | 0.36 | 0.88 |
| SOL & TA | *r* | 0.46 | **0.59** | 0.43 | 0.50 | 0.33 | 0.31 | 0.37 | 0.40 | 0.46 | -0.32 |
|  | *p* | 0.06 | **0.03** | 0.16 | 0.10 | 0.19 | 0.24 | 0.20 | 0.20 | 0.30 | 0.50 |

Spearman correlation coefficient (r) and p-values. Significant associations are indicated in bold.
L1-L6 indicate perturbation level. LG = lateral gastrocnemius; MG = medial gastrocnemius; SOL = soleus; TA= tibialis anterior.

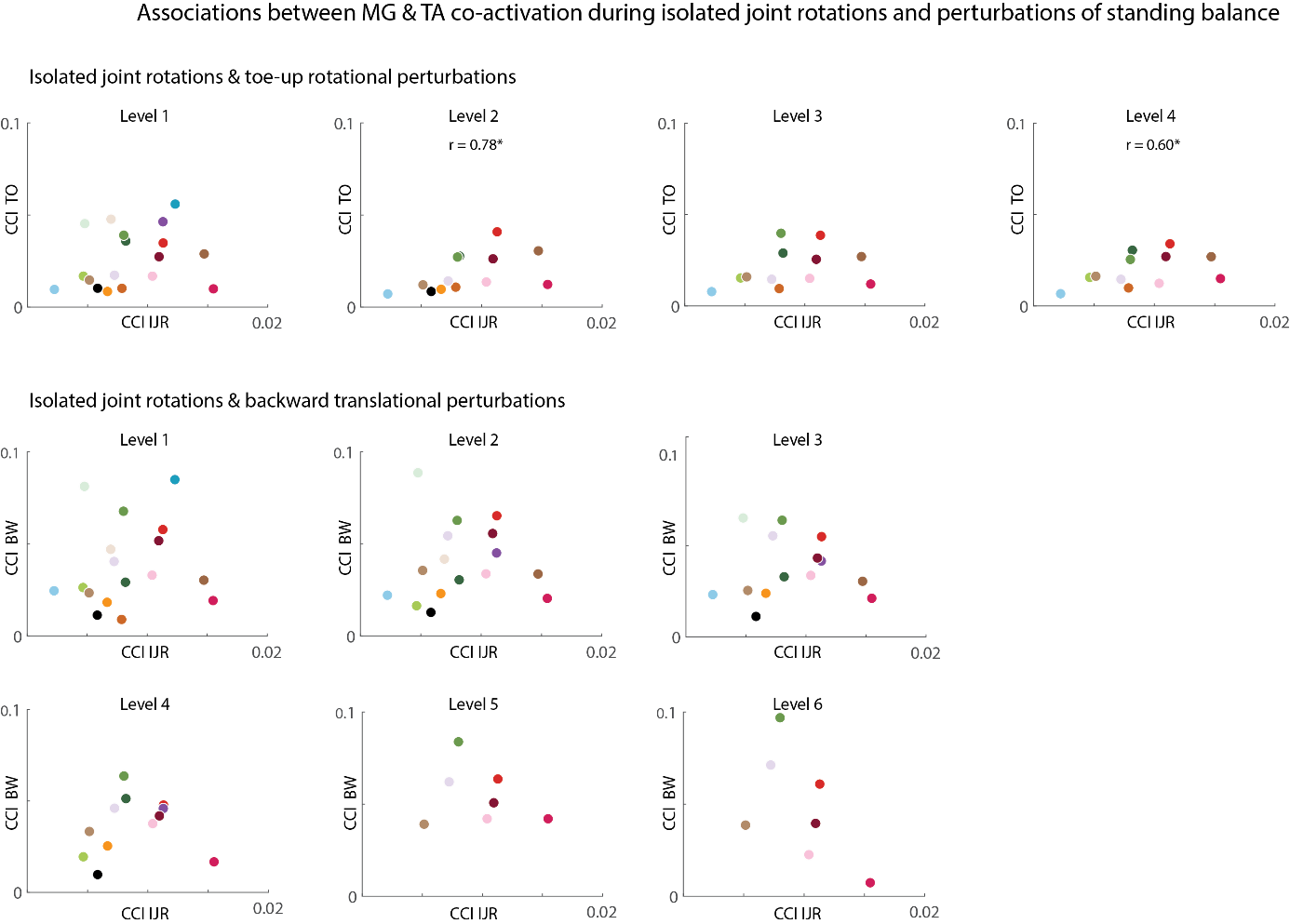
Figure S4: Associations between medial gastrocnemius (MG) and tibialis anterior (TA) co-activation (CCI) during isolated joint rotations (IJR) and perturbations of standing balance (toe-up rotations (TO): upper part; backward translations (BW): lower part) for each level. Each color represents one child. Significant associations (p<0.05) are indicated with a star and corresponding r value.

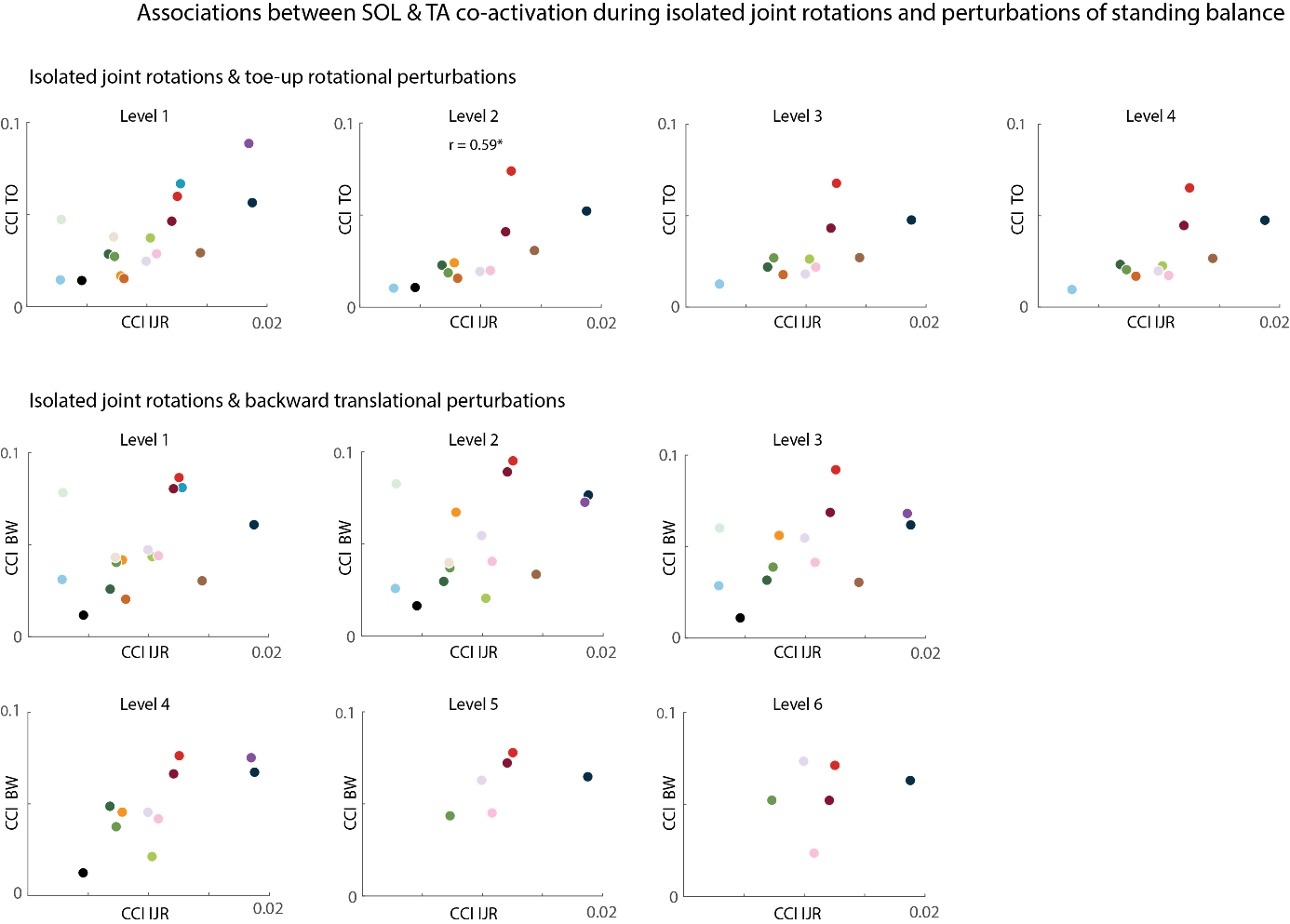
Figure S5: Associations between soleus (SOL) and tibialis anterior (TA) co-activation (CCI) during isolated joint rotations (IJR) and perturbations of standing balance (toe-up rotations (TO): upper part; backward translations (BW): lower part) for each level. Each color represents one child. Significant associations (p<0.05) are indicated with a star and corresponding r value.

**S5. Mixed linear model for muscle co-activation during isolated joint rotations and perturbations of standing balance**

Table S5: Statistical outcome parameters for the relation between muscle co-activation during isolated joint rotations and perturbations of standing balance.

| **CCI** | **p-value** | | |
| --- | --- | --- | --- |
|  | **Level** | **IJR** | **Level:IJR** |
|  | Toe-up rotations | | |
| LG & TA | 0.426 | **0.006** | 0.523 |
| MG & TA | 0.296 | **0.003** | 0.113 |
| SOL & TA | 0.177 | **0.047** | 0.513 |
|  | Backward translations | | |
| LG & TA | 0.144 | **0.004** | **0.025** |
| MG & TA | **0.002** | **0.024** | **0.001** |
| SOL & TA | 0.618 | 0.272 | 0.213 |

Significant differences are indicated in bold.
CCI = co-contraction index; IJR = isolated joint rotation; LG = lateral gastrocnemius; MG = medial gastrocnemius; SOL = soleus; TA = tibialis anterior.

**S6. Associations for the mean muscle activity during isolated joint rotations and perturbations of standing balance.**

Table S6: Associations (r and p-values) for mean muscle activity during isolated joint rotations and perturbations of standing balance.

|  |  | **Toe-up rotations** | | | | **Backward translations** | | | | | |
| --- | --- | --- | --- | --- | --- | --- | --- | --- | --- | --- | --- |
|  |  | **L1** | **L2** | **L3** | **L4** | **L1** | **L2** | **L3** | **L4** | **L5** | **L6** |
| LG | *r* | 0.11 | 0.40 | 0.16 | 0.25 | 0.35 | 0.36 | 0.21 | 0.24 | 0.48 | -0.02 |
|  | *p* | 0.66 | 0.14 | 0.57 | 0.36 | 0.14 | 0.14 | 0.44 | 0.41 | 0.16 | 0.96 |
| MG | *r* | 0.31 | 0.31 | 0.36 | 0.37 | 0.00 | 0.13 | 0.28 | 0.21 | 0.44 | 0.27 |
|  | *p* | 0.18 | 0.27 | 0.19 | 0.17 | 0.99 | 0.61 | 0.29 | 0.46 | 0.20 | 0.45 |
| SOL | *r* | **0.47** | **0.74** | **0.57** | **0.72** | 0.37 | 0.36 | 0.32 | 0.41 | 0.43 | 0.02 |
|  | *p* | **0.04** | **0.002** | **0.03** | **0.004** | 0.13 | 0.16 | 0.24 | 0.17 | 0.25 | 0.98 |
| TA | *r* | 0.26 | 0.47 | 0.06 | 0.37 | 0.45 | 0.28 | 0.35 | 0.23 | -0.48 | -0.36 |
|  | *p* | 0.28 | 0.09 | 0.84 | 0.21 | 0.06 | 0.28 | 0.20 | 0.45 | 0.24 | 0.39 |

Spearman correlation coefficient (r) and p-values (p)
L1-L6 indicate perturbation level. Significant values are indicated in bold.
LG = lateral gastrocnemius; MG= medial gastrocnemius; SOL = soleus; TA = tibialis anterior;

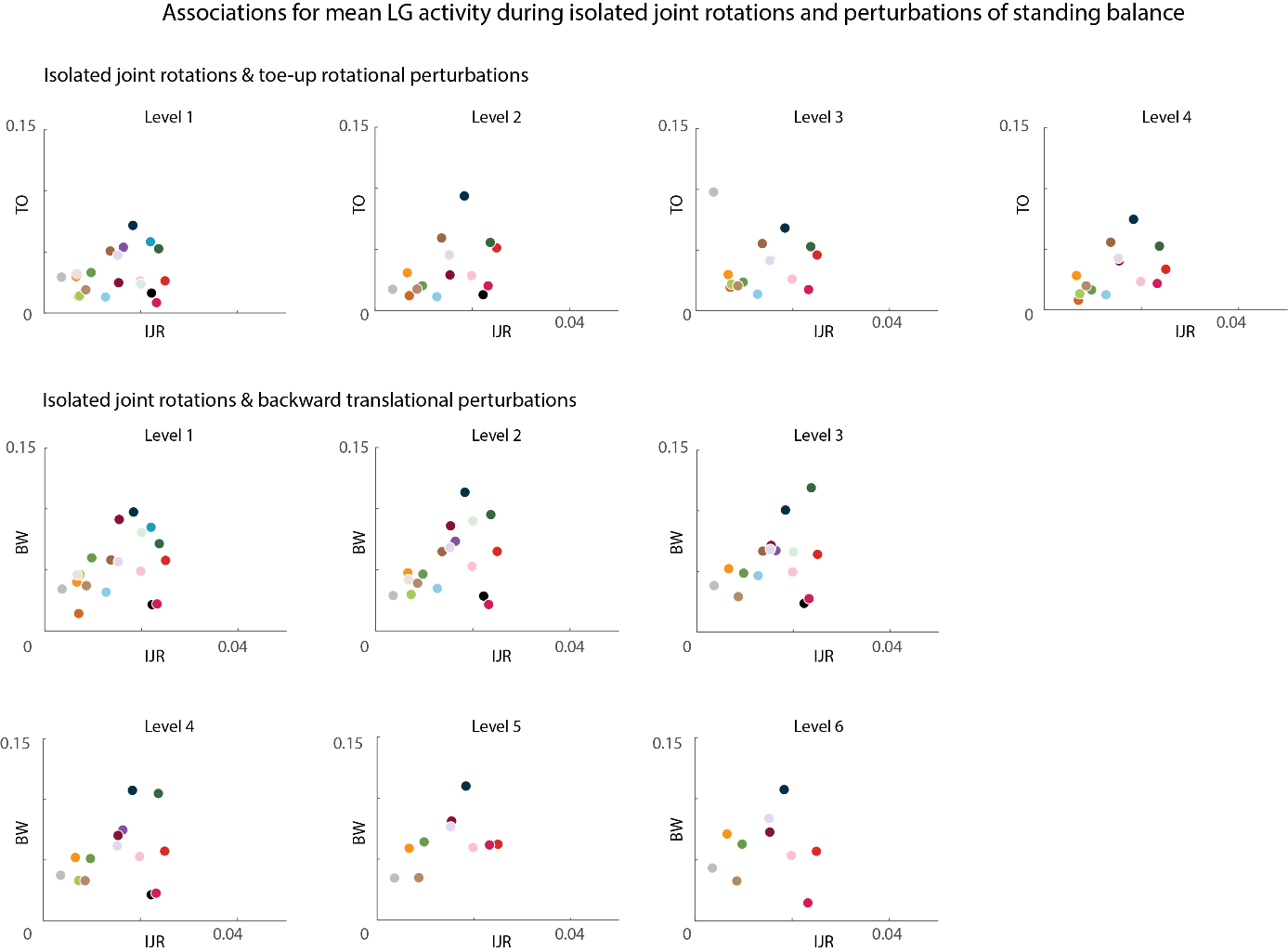

Figure S6: Associations for lateral gastrocnemius (LG) mean muscle activity during isolated joint rotations (IJR) and perturbations of standing balance (toe-up rotations (TO): upper part; backward translations (BW): lower part) for each level. Each color represents one child. No associations were significant.

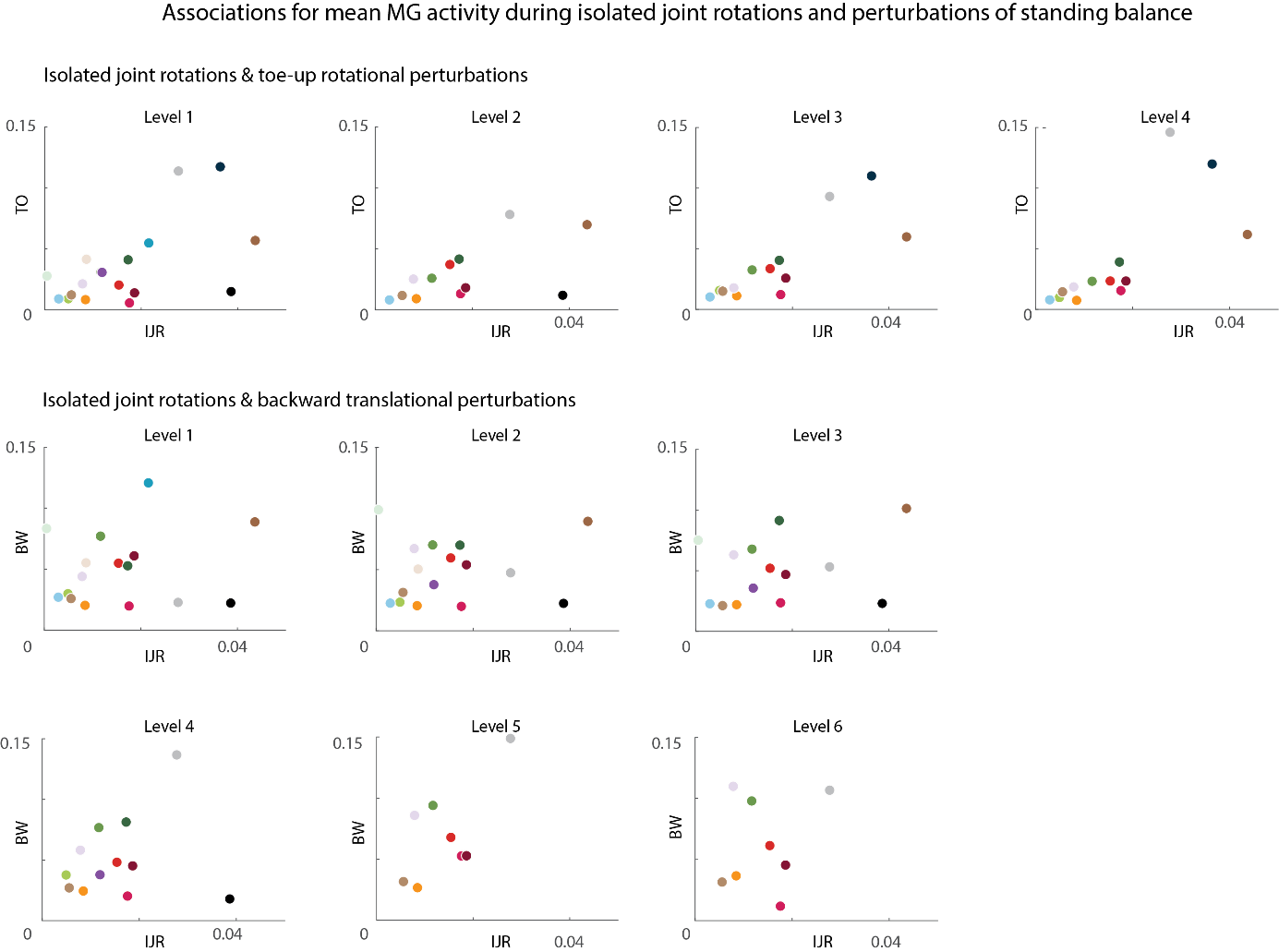

Figure S7: Associations for medial gastrocnemius (MG) mean muscle activity during isolated joint rotations (IJR) and perturbations of standing balance (toe-up rotations (TO): upper part; backward translations (BW): lower part) for each level. Each color represents one child. No associations were significant.

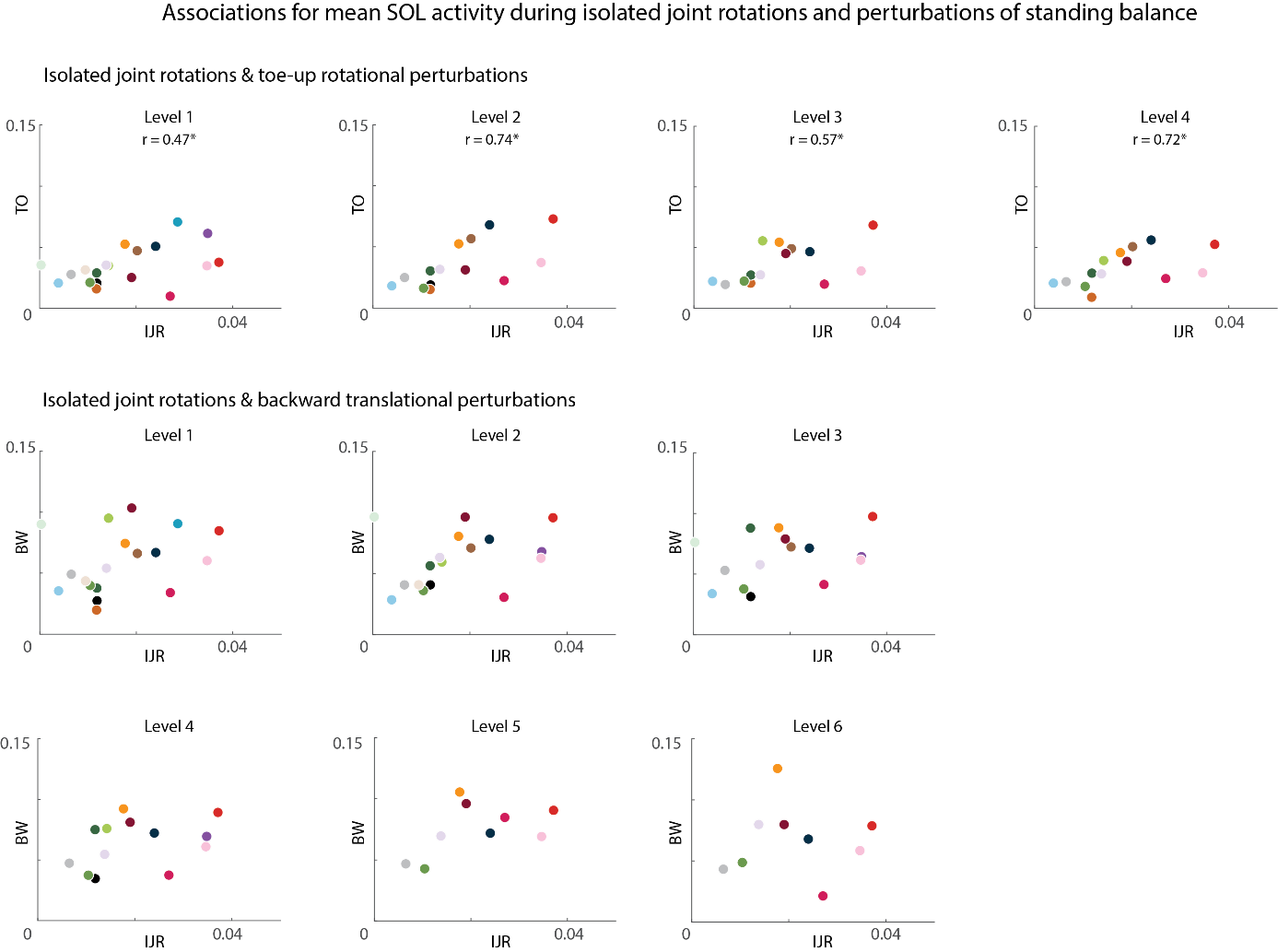

Figure S8: Associations for soleus (SOL) mean muscle activity during isolated joint rotations (IJR) and perturbations of standing balance (toe-up rotations (TO): upper part; backward translations (BW): lower part) for each level. Each color represents one child. Significant associations are indicated with spearman correlation coefficient (r) and a star.

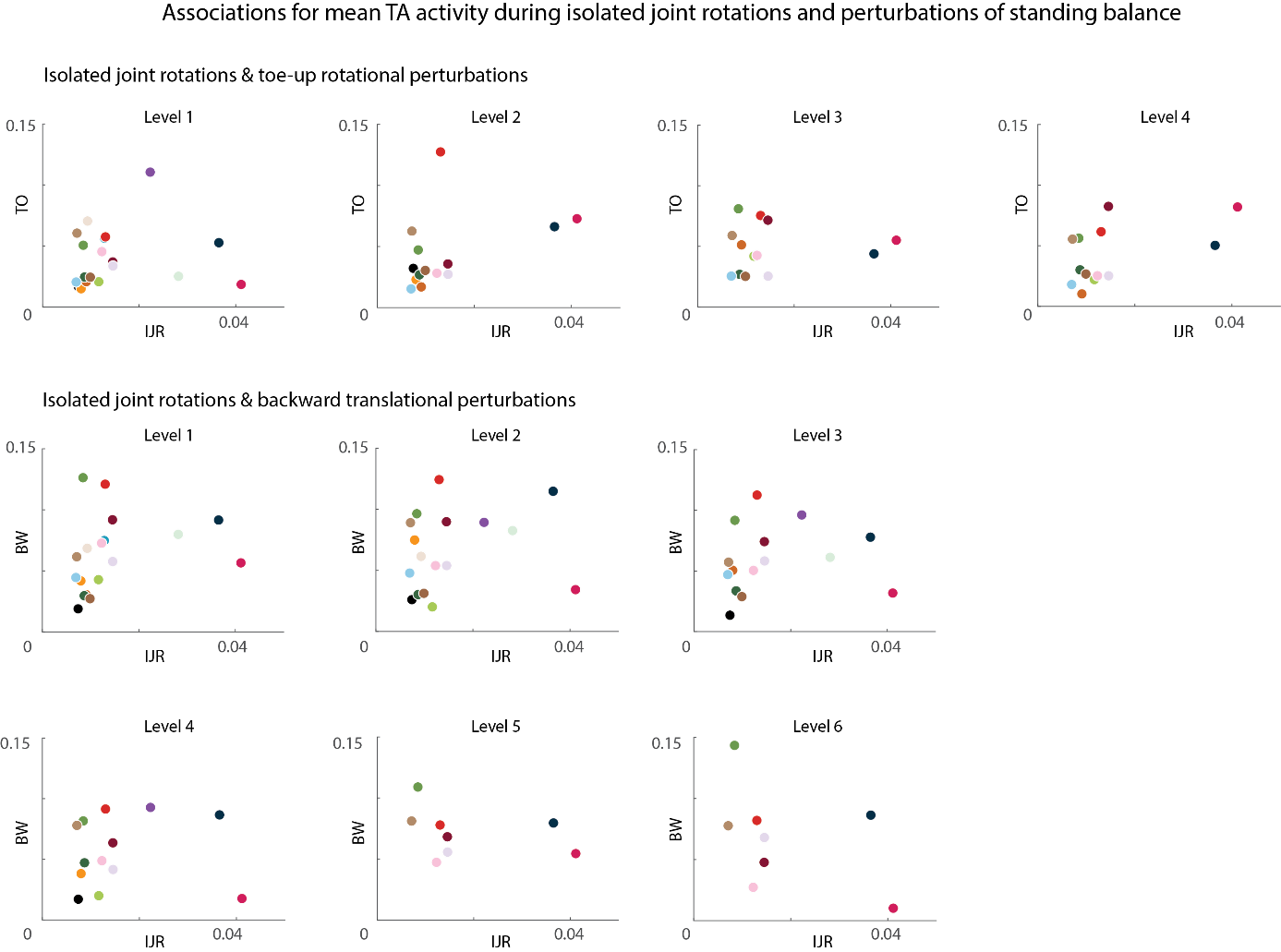

Figure S9: Associations for tibialis anterior (TA) mean muscle activity during isolated joint rotations (IJR) and perturbations of standing balance (toe-up rotations (TO): upper part; backward translations (BW): lower part) for each level. Each color represents one child. No associations were significant.

**S7. Mixed linear model for mean muscle activity during isolated joint rotations and perturbations of standing balance**

Table S7: Statistical outcome parameters for the relation between mean muscle activity during isolated joint rotations and perturbations of standing balance.

| **Mean muscle activity** | **p-value** | | |
| --- | --- | --- | --- |
|  | **Level** | **IJR** | **Level:IJR** |
|  | Toe-up rotations | | |
| LG | **0.013** | 0.472 | **0.033** |
| MG | 0.921 | 0.695 | 0.942 |
| SOL | 0.166 | **0.034** | **0.005** |
| TA | 0.143 | 0.696 | 0.058 |
|  | Backward translations | | |
| LG | 0.088 | 0.106 | **0.028** |
| MG | 0.176 | 0.796 | 0.777 |
| SOL | 0.442 | 0.166 | 0.440 |
| TA | 0.820 | 0.184 | 0.351 |

Significant differences are indicated in bold.
IJR = isolated joint rotation; LG = lateral gastrocnemius; MG = medial gastrocnemius; SOL = soleus; TA = tibialis anterior.

**S8. Associations and mixed linear model for center of mass excursion during perturbations of standing balance and muscle co-activation during isolated joint rotations**

Table S8: Associations (Spearman r and p-values) between center of mass excursion during perturbations of standing balance and muscle co-activation during isolated joint rotations.

|  |  | **Toe-up rotations** | | | | **Backward translations** | | | | | |
| --- | --- | --- | --- | --- | --- | --- | --- | --- | --- | --- | --- |
|  |  | **L1** | **L2** | **L3** | **L4** | **L1** | **L2** | **L3** | **L4** | **L5** | **L6** |
| LG & TA | *r* | 0.15 | **0.55** | 0.06 | -0.06 | 0.33 | 0.11 | 0.36 | -0.09 | 0.32 | 0.37 |
|  | *p* | 0.53 | **0.04** | 0.83 | 0.84 | 0.16 | 0.67 | 0.16 | 0.75 | 0.37 | 0.29 |
| MG & TA | *r* | 0.15 | 0.48 | 0.06 | 0.14 | 0.11 | 0.01 | 0.01 | -0.16 | 0.18 | 0.20 |
|  | *p* | 0.54 | 0.07 | 0.83 | 0.61 | 0.64 | 0.97 | 0.97 | 0.59 | 0.63 | 0.58 |
| SOL & TA | *r* | 0.04 | 0.29 | 0.02 | -0.30 | 0.20 | 0.01 | -0.04 | -0.24 | 0.30 | 0.32 |
|  | *p* | 0.86 | 0.31 | 0.95 | 0.30 | 0.42 | 0.97 | 0.88 | 0.44 | 0.44 | 0.41 |

Significant differences are indicated in bold. L1-L6 indicate perturbation levels.
LG = lateral gastrocnemius; MG = medial gastrocnemius; SOL = soleus; TA = tibialis anterior.

Table S9: Statistical outcome parameters for the relation between center of mass excursion and muscle co-activation during isolated joint rotations.

| **CoM excursion** | **p-value** | | |
| --- | --- | --- | --- |
|  | **Level** | **IJR** | **Level:IJR** |
|  | Toe-up rotations | | |
| LG & TA | **0.001** | 0.988 | 0.286 |
| MG & TA | **<0.001** | 0.860 | 0.347 |
| SOL & TA | **<0.001** | 0.728 | 0.265 |
|  | Backward translations | | |
| LG & TA | **<0.001** | 0.584 | 0.936 |
| MG & TA | **<0.001** | 0.902 | 0.685 |
| SOL & TA | **<0.001** | 0.738 | 0.747 |

Significant differences are indicated in bold.
CoM = center of mass; IJR = Isolated joint rotation; LG = lateral gastrocnemius; MG = medial gastrocnemius; SOL = soleus; TA = tibialis anterior.

**S9. Associations and mixed linear model for Modified Ashworth Score and muscle co-activation during perturbations of standing balance**

Table S10: Associations (Spearman r and p-values) between MAS and muscle co-activation during perturbations of standing balance.

|  | **Toe-up rotations** | | | | **Backward translations** | | | | | |
| --- | --- | --- | --- | --- | --- | --- | --- | --- | --- | --- |
| *R* | **L1** | **L2** | **L3** | **L4** | **L1** | **L2** | **L3** | **L4** | **L5** | **L6** |
| LG & TA | 0.09 | 0.17 | 0.11 | 0.16 | 0.06 | 0.45 | **0.65** | **0.58** | **0.89** | 0.73 |
| MG & TA | 0.08 | 0.19 | 0.01 | 0.04 | 0.09 | **0.52** | 0.51 | 0.56 | 0.7 | 0.54 |
| SOL & TA | -0.05 | 0.13 | 0.06 | 0.05 | 0.07 | **0.52** | 0.51 | 0.55 | 0.66 | **0.84** |
| *p-value* |  |  |  |  |  |  |  |  |  |  |
| LG & TA | 0.71 | 0.57 | 0.71 | 0.60 | 0.82 | 0.07 | **0.01** | **0.04** | **0.005** | 0.05 |
| MG & TA | 0.75 | 0.51 | 0.98 | 0.89 | 0.72 | **0.03** | 0.05 | 0.05 | 0.06 | 0.17 |
| SOL & TA | 0.83 | 0.67 | 0.85 | 0.89 | 0.79 | **0.04** | 0.06 | 0.06 | 0.13 | **0.04** |

Significant differences are indicated in bold. L1-L6 indicate perturbation levels.
LG = lateral gastrocnemius; MG = medial gastrocnemius; SOL = soleus; TA = tibialis anterior.

Table S11: Statistical outcome parameters for the relation between MAS and muscle co-activation during perturbations of standing balance.

| **CCI** | **p-value** | | |
| --- | --- | --- | --- |
|  | **Level** | **MAS** | **Level:MAS** |
|  | Toe-up rotations | | |
| LG & TA | 0.901 | 0.604 | 0.169 |
| MG & TA | 0.972 | 0.193 | 0.152 |
| SOL & TA | **0.012** | 0.820 | **0.016** |
|  | Backward translations | | |
| LG & TA | 0.946 | 0.736 | 0.353 |
| MG & TA | 0.821 | 0.385 | 0.547 |
| SOL & TA |  |  |  |

Significant differences are indicated in bold.
CCI = co-contraction index; MAS = Modified Ashworth Score; LG = lateral gastrocnemius; MG = medial gastrocnemius; SOL = soleus; TA = tibialis anterior.

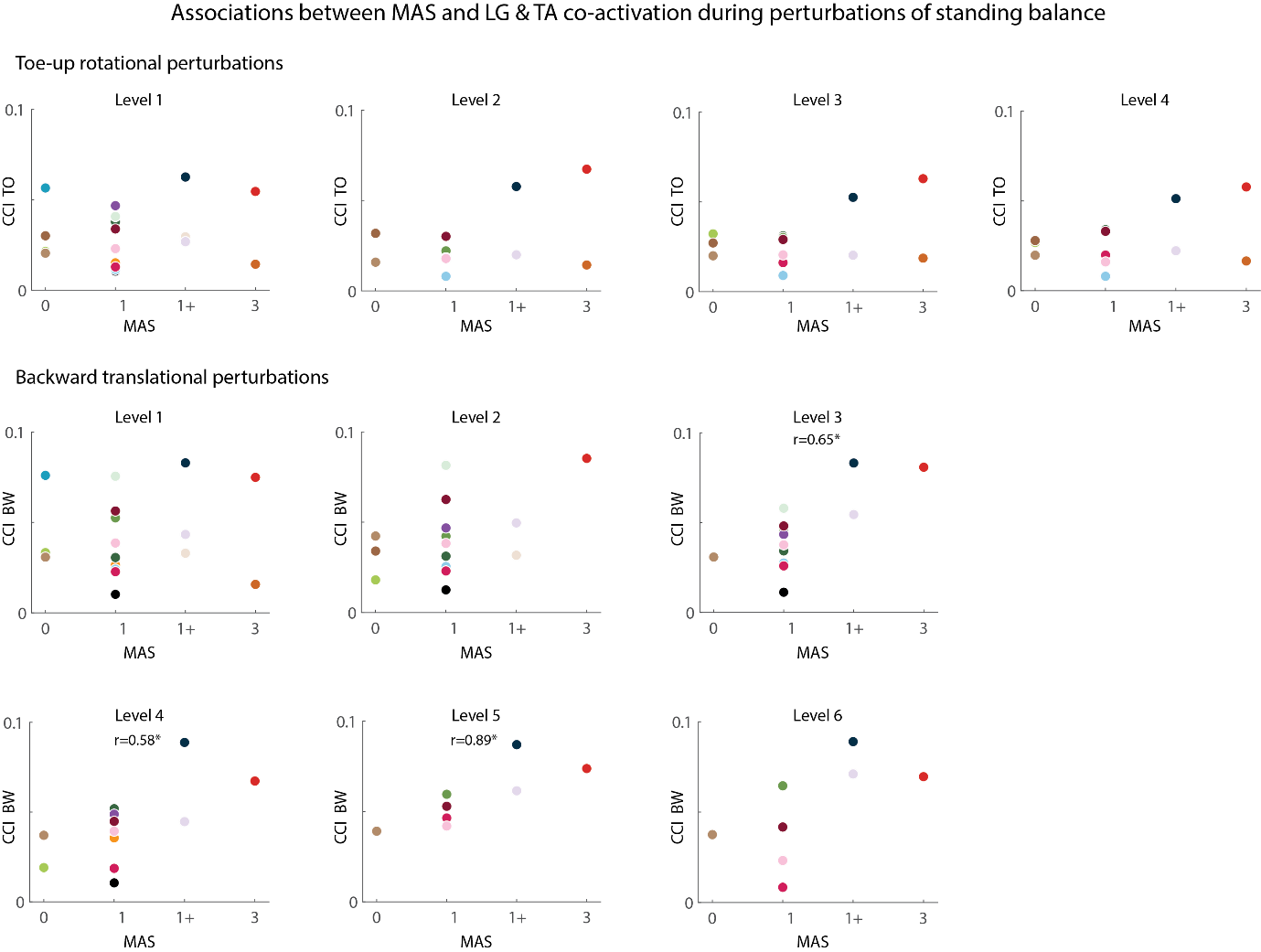
Figure S10: Associations between MAS and muscle co-activation (CCI) between lateral gastrocnemius (LG) and tibialis anterior (TA) during perturbations of standing balance (toe-up rotations (TO): upper part; backward translations (BW): lower part) for each level. Each color represents one child. Significant associations (p<0.05) are indicated with a star and corresponding r value.

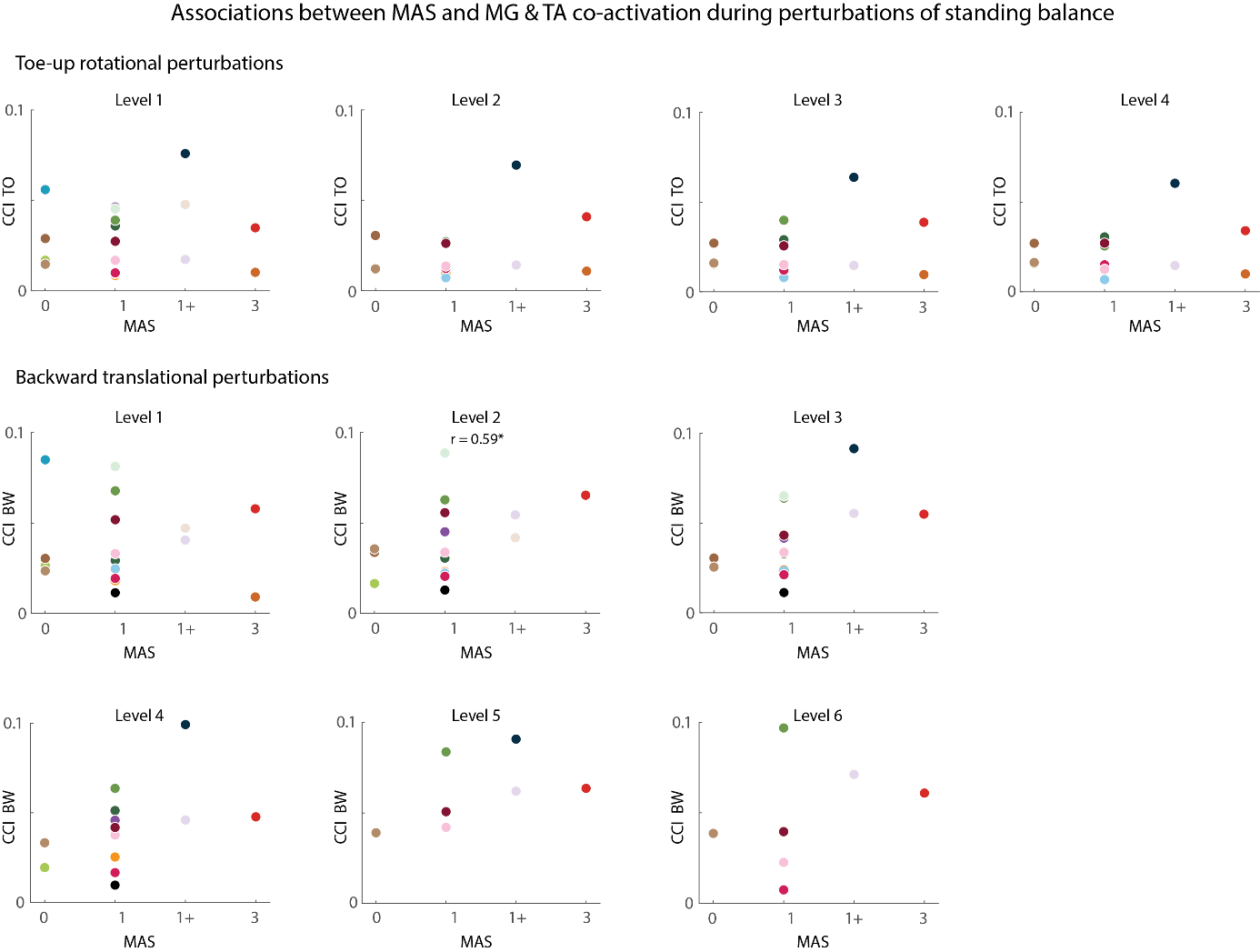
Figure S11: Associations between MAS and muscle co-activation (CCI) between medial gastrocnemius (MG) and tibialis anterior (TA) during perturbations of standing balance (toe-up rotations (TO): upper part; backward translations (BW): lower part) for each level. Each color represents one child. Significant associations (p<0.05) are indicated with a star and corresponding r value.

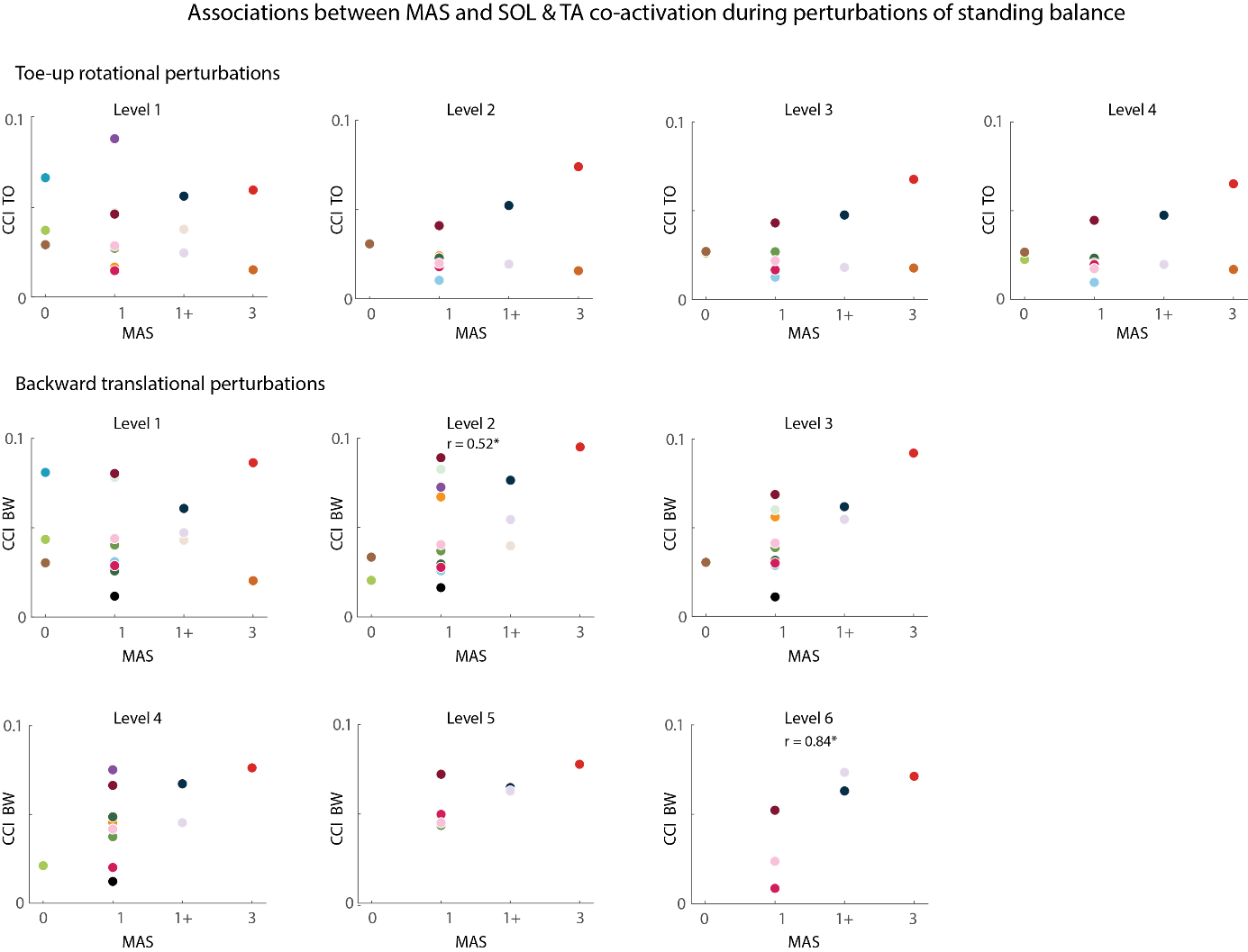

Figure S12: Associations between MAS and muscle co-activation (CCI) between soleus (SOL) and tibialis anterior (TA) during perturbations of standing balance (toe-up rotations (TO): upper part; backward translations (BW): lower part) for each level. Each color represents one child. Significant associations (p<0.05) are indicated with a star and corresponding r value.

**S10. Associations between passive joint stiffness and muscle co-activation during perturbations of standing balance**

We calculated the inverse dynamic torque when the ankle angle was zero degrees dorsiflexion during a slow isolated joint rotation. During a slow isolated joint rotation, the examiner rotates the joint from plantarflexion towards end-range of motion in dorsiflexion during 5 sec.

Table S12: Associations (Spearman r and p-values) between passive muscle stiffness and muscle co-activation during perturbations of standing balance.

|  |  | **Toe-up rotations** | | | | **Backward translations** | | | | | |
| --- | --- | --- | --- | --- | --- | --- | --- | --- | --- | --- | --- |
|  |  | **L1** | **L2** | **L3** | **L4** | **L1** | **L2** | **L3** | **L4** | **L5** | **L6** |
| LG & TA | *r* | 0.01 | -0.01 | -0.13 | -0.10 | 0.07 | -0.23 | -0.10 | -0.28 | -0.31 | -0.43 |
|  | *p* | 0.96 | 0.97 | 0.70 | 0.75 | 0.80 | 0.43 | 0.75 | 0.40 | 0.46 | 0.30 |
| MG & TA | *r* | 0.02 | 0.01 | -0.27 | -0.24 | 0.10 | -0.21 | 0.03 | -0.29 | -0.31 | -0.45 |
|  | *p* | 0.93 | 0.99 | 0.39 | 0.46 | 0.70 | 0.47 | 0.93 | 0.39 | 0.46 | 0.27 |
| SOL & TA | *r* | 0.34 | 0.15 | 0.02 | -0.25 | -0.20 | 0.01 | -0.24 | -0.12 | -0.25 | -0.36 |
|  | *p* | 0.58 | 0.95 | 0.45 | 0.56 | 0.98 | 0.44 | 0.71 | 0.49 | 0.44 | 0.09 |

No significant associations were found. L1-L6 indicate perturbation levels. LG = lateral gastrocnemius; MG = medial gastrocnemius; SOL = soleus; TA = tibialis anterior.

**S11. Mean and range of CCI and mean muscle activity for typically developing children.**

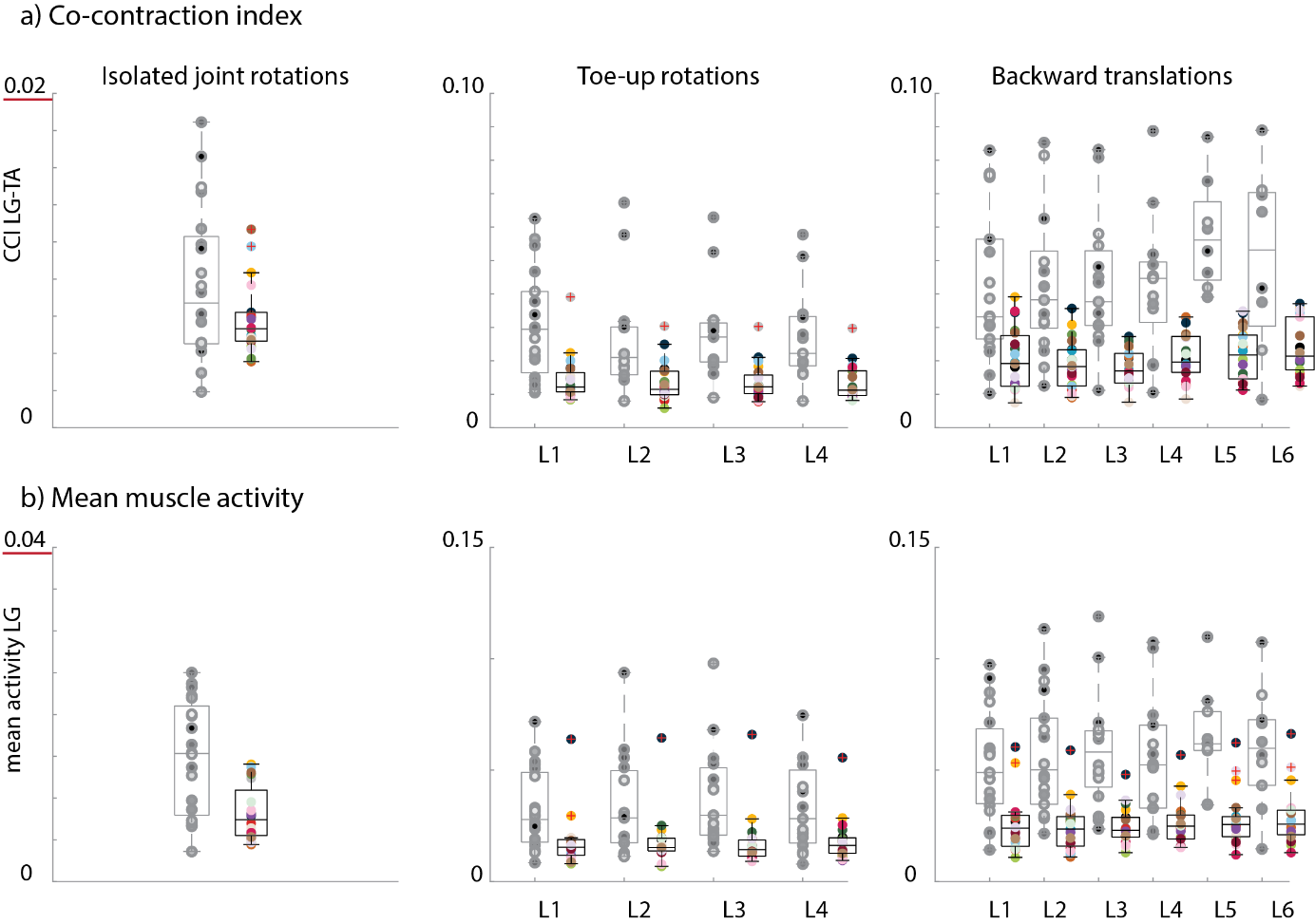

Figure S13: Co-contraction index for LG-TA **(a)** and mean muscle activity for LG **(b)** across all typically developing children (in color) for isolated joint rotations (left column), toe-up rotations (middle column), and backward translations (right column). Every dot represents one child. Data of children with CP is indicated in grey as reference. L1-L6 = Levels 1 to 6.

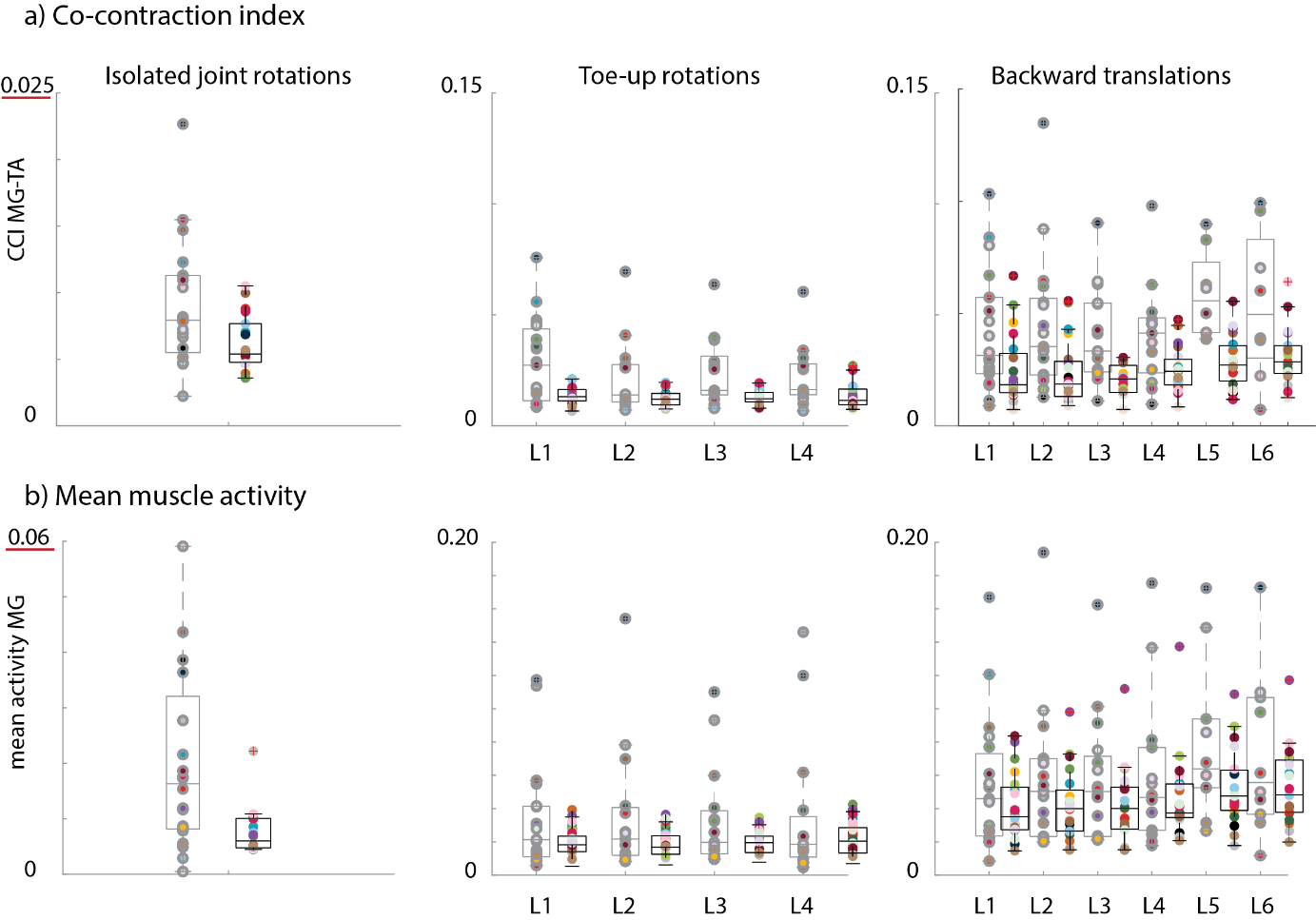

Figure S14: Co-contraction index for MG-TA **(a)** and mean muscle activity for MG **(b)** across all typically developing children (in color) for isolated joint rotations (left column), toe-up rotations (middle column), and backward translations (right column). Every dot represents one child. Data of children with CP is indicated in grey as reference. L1-L6 = Levels 1 to 6.

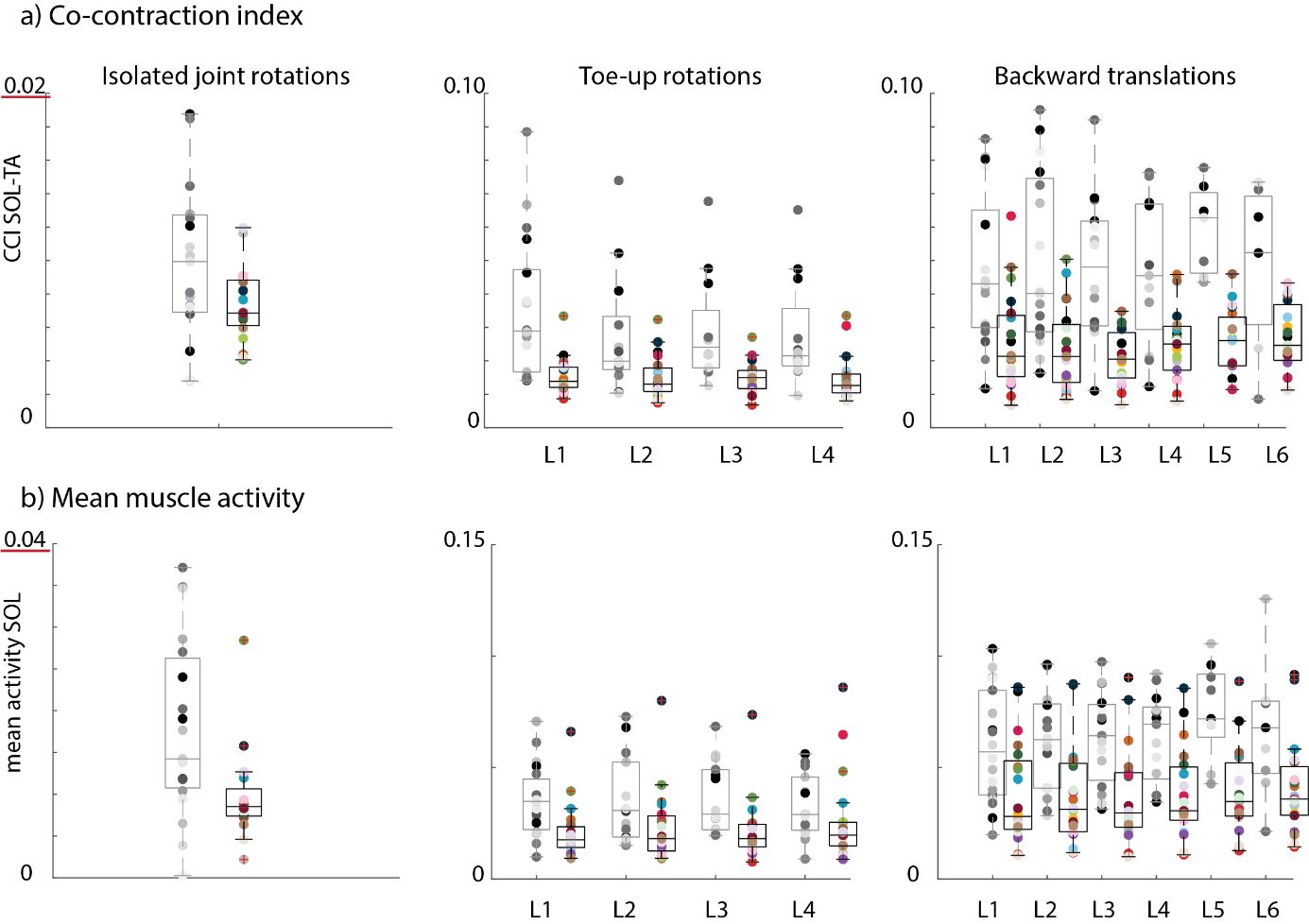

Figure S15: Co-contraction index for SOL-TA **(a)** and mean muscle activity for SOL **(b)** across all typically developing children (in color) for isolated joint rotations (left column), toe-up rotations (middle column), and backward translations (right column). Every dot represents one child. Data of children with CP is indicated in grey as reference. L1-L6 = Levels 1 to 6.
